# Evaluation of a Potential Relationship between CFTR Modulators, CFTR function and Neuropsychiatric Symptoms in People with CF

**DOI:** 10.1101/2025.10.16.25338186

**Authors:** Fredrick Van Goor, Laurent Francioli, Patrick R. Sosnay, James L. Kreindler, Neil Ahluwalia, Mark J. Daly, Weichao Chen, David Altshuler

## Abstract

**Rationale:** Cystic fibrosis (CF) is a chronic, life-limiting genetic disease. Treatment with CFTR modulators (CFTRm), such as the triple combination therapies of elexacaftor (ELX)/tezacaftor (TEZ)/ivacaftor (IVA) and vanzacaftor (VNZ), TEZ, and deutivacaftor (D-IVA), have significantly improved clinical outcomes and quality of life for many people with CF. While most individuals treated with CFTRm report no change or improvement in neuropsychiatric symptoms, a minority have reported new or worsening symptoms. This has led to hypotheses that CFTRm or CFTR function could have relationships to neuropsychiatric symptoms.

**Objectives:** Test specific hypotheses about potential relationships between CFTRm, CFTR function, and neuropsychiatric symptoms.

**Methods:** To evaluate potential off-target or on-target effects of CFTRm and/or CFTR function on neuropsychiatric symptoms, we conducted four independent analyses: (a) *in vitro* off-target screening at clinically-relevant free concentrations; (b) nonclinical, *in vivo* behavioral pharmacology studies; (c) analysis of clinical trial and real-world evidence of CFTRm neuropsychiatric adverse events; and (d) human genetic analysis of *CFTR* functional variants in ∼1 million individuals diagnosed with neuropsychiatric disorders.

**Measurements and Main Results:** No off-target effects of ELX, VNZ, TEZ, or IVA were observed *in vitro* at clinically-relevant free concentrations, nor were any behavioral findings observed *in vivo* animal studies. Clinical trial and real-world data covering >250,000 patient-years of experience with CFTRm did not show any increase in risk of neuropsychiatric adverse events. Human genetic analyses of functional *CFTR* variants did not identify association with any of 23 neuropsychiatric conditions.

**Conclusions:** Multiple independent lines of investigation failed to show any evidence for off-target or on-target effects of CFTRm and/or *CFTR* function on neuropsychiatric symptoms.

## Introduction

Cystic fibrosis (CF) is a chronic, life-limiting genetic disease that is progressive from birth, leading to multi-organ damage and early death.(1) As with other serious chronic diseases, people with CF (pwCF) have an increased risk for adverse psychological symptoms including rates of depression and anxiety that are higher than those of comparable healthy individuals.(2, 3) A 2014 study measured rates of depression and anxiety in 6,088 pwCF across nine countries prior to the widespread introduction of CFTR modulators (CFTRm): symptoms of depression were reported in 5% to 19% of adolescents and 13% to 29% of adults, while symptoms of anxiety were observed in 22% of adolescents and 32% of adults with CF.(3)

CFTRm therapies that directly treat the underlying cause of CF have altered the disease trajectory for many pwCF.(4) Triple combinations of elexacaftor (ELX), tezacaftor (TEZ), ivacaftor (IVA) and vanzacaftor (VNZ), TEZ, and deutivacaftor (D-IVA) improve lung function and quality of life, reduce pulmonary exacerbations, hospitalizations and lung transplants, and may significantly extend life expectancy.(5–7)

Prior analysis of clinical trial and real-world evidence documented that most pwCF treated with ELX/TEZ/IVA experienced no change or an improvement in depression-related events.(8) However, a minority of people treated with CFTRm have reported new or worsening neuropsychological symptoms including depression, anxiety, sleep disturbance, and cognitive changes such as brain fog.(9, 10) These symptoms typically occur within the first three months of treatment and, in some cases, are associated with treatment interruption or discontinuation.(9) These reports have led to hypotheses regarding a potential role for either off-target pharmacological effects of CFTRm and/or of CFTR function in the brain on neuropsychiatric traits. For example, published *in vitro* experiments reported binding of CFTRm at free (unbound) concentrations far exceeding clinically relevant free exposures, leading to the hypothesis that CFTRm may exert off-target effects on central nervous system receptors or transmitters associated with neuropsychiatric symptoms.(11, 12) It has also been hypothesized that alterations in function of CFTR expressed in the brain could potentially contribute to mood and cognitive changes.(12, 13)

To further evaluate these hypotheses, we: 1) tested *in vitro* off-target binding activity at clinically relevant free concentrations of ELX, VNZ, TEZ, and IVA across a broad panel of neuropsychiatric targets, 2) assessed neurobehavioral endpoints in rats following oral administration of ELX, VNZ, TEZ, or IVA, 3) searched for association between CFTRm and neuropsychiatric observations across clinical trials and real-world evidence studies, and (4) analyzed functional CFTR variants for potential associations in ∼1 million people diagnosed with neuropsychiatric diseases. Collectively, these studies did not identify any evidence for off-target or on-target effects of CFTRm or CFTR function on neuropsychiatric traits.

## Methods

### Receptor binding assays

Off-target binding activity of CFTRm and major circulating metabolites was tested using the LeadProfilingScreen and/or SpectrumScreen® Panel methods(14) on a comprehensive panel of neuropsychiatric targets.

### Rat neurobehavioral studies

Sprague Dawley rats were randomly assigned to receive ELX, VNZ, TEZ, or IVA by oral gavage at three dose levels, with vehicle controls included in all studies and a positive control (chlorpromazine) in the IVA study. Neurobehavioral effects were assessed using the Irwin test (IVA) or Functional Observational Battery (FOB; ELX, VNZ, and TEZ) at multiple time points post-dose to capture peak plasma concentrations (C_max_).(15, 16) Parameters evaluated included activity, autonomic function, neuromuscular and sensorimotor responses, and general health (**Table E1**). Data were compared between treated and control groups to identify any neurobehavioral changes, with statistical analysis performed for FOB endpoints. Detailed methods and analysis are provided in the Appendix.

### Determination of the free concentration at the C_max_ for ELX, VNZ, TEZ, and IVA

Free concentrations of ELX, VNZ, TEZ, and IVA at the C_max_ were determined by multiplying total C_max_ by the free fraction (unbound) as determined using the equilibrium dialysis method (see Appendix). Standard noncompartmental analyses were used to determine pharmacokinetic (PK) parameters such as C_max._ Population PK methods were used to characterize exposures of ELX, VNZ, TEZ, and IVA in healthy subjects and CF participants incorporating the effects of demographic characteristics and other covariates on PK.

### Rat brain distribution studies

The tissue distribution of CFTRm was determined by administering radiolabeled ^14^C to male Sprague-Dawley rats and measuring tissue specific levels at various timepoints postdosing (see Appendix).

### Pooled CFTRm clinical trials

Relevant psychiatric events from the pivotal, placebo-controlled and open-label extension trials with CFTRm ELX/TEZ/IVA, TEZ/IVA, LUM/IVA, and IVA were identified by preferred term. Additionally, relevant events from over 60 Phase 2 and 3, controlled and uncontrolled clinical trials in with ELX/TEZ/IVA, TEZ/IVA, LUM/IVA, and IVA were identified and pooled for comparison with pooled placebo. The data are summarized for events, serious events, and events leading to treatment discontinuation. Phase 3 VNZ/TEZ/D-IVA trial data on neuropsychiatric side effects was reported in Keating et al. (7)

### Real-world outcomes from US and German CF registries

Prevalence of depression and anxiety diagnosis in pwCF treated with ELX/TEZ/IVA was assessed using United States (US) and German CF registries as part of an ongoing post-approval safety study. Data from 8,967 US and 1,028 German patients who started ELX/TEZ/IVA between late 2019 and 2021 and had non-missing data in all years were analyzed, comparing prevalence over 4 years post-treatment to the 5 years before treatment initiation.

### Systematic Review of Published Literature

A systematic literature review (SLR) was conducted to evaluate observational studies assessing neuropsychiatric outcomes before and after ELX/TEZ/IVA initiation (**Figure E1**). The SLR adhered to established methods for conducting systematic reviews and were reported in accordance with the Preferred Reporting Items for Systematic Literature Reviews and Meta-Analyses (PRISMA) statement. (17)

**Figure 1:**
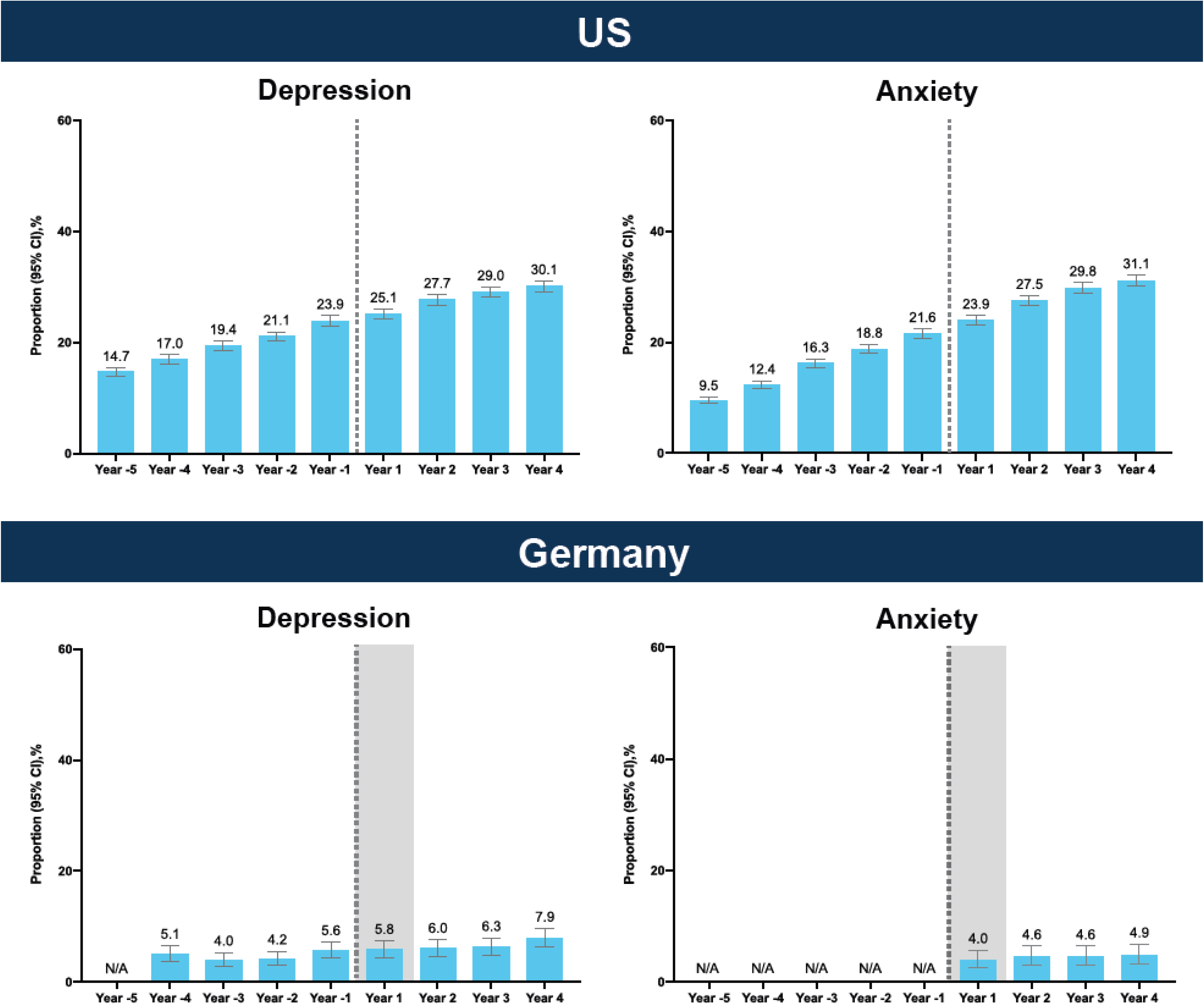
Real World registry diagnoses of depression and anxiety pre- and post-ELX/TEZ/IVA initiation. Proportion of diagnosed depression (left panels) and anxiety (right panels) in people with cystic fibrosis (pwCF) before and after initiation of ELX/TEZ/IVA therapy. Data are shown for five years prior to and four years following treatment initiation, using US (top panel) and German (bottom) CF patient registries. Analyses included 8,967 patients in the US and 1,028 patients in Germany with non-missing data in all years; number of patients with non-missing data may vary across variables. Shaded area in Germany includes a mixture of pre-treatment and post-treatment data. The vertical dashed line indicates the time of ELX/TEZ/IVA initiation. Years in which these diagnoses were not collected in the registry are labelled N/A. Anxiety diagnoses were not captured by the German CF Registry in the pre-treatment period.

### Genetic studies

To assess whether CFTR modulation in the brain is associated with neuropsychiatric traits, we analyzed the common CF-causing *F508del*-CFTR variant as well as that of rare, severe *CFTR* premature termination variants (Class I variants).

First, we evaluated association data in a broad and unbiased set of phenotypes using publicly available data from multiple biobanks. For *F508del*-CFTR associations, we used data from the UKBB/FinnGen browser (18) that provides meta-analysis of genome-wide associations studies (GWAS) results from two large biobanks (UK Biobank and FinnGen) encompassing 873 phenotypes in approximately 1 million individuals. For CFTR protein-truncating variants, we used burden tests associations from Genebass (19) spanning 4,529 phenotypes across 394,841 individuals from the UK biobank, and All by all (20) encompassing ∼3,400 traits across ∼250,000 individuals from the All of Us biobank. We searched for potential associations across all phenotypes. The pre-specified threshold for genome-wide association is 10^-8^; threshold for suggestive association in a phenome-wide scan is p <10^-4^ for exome sequencing, and <10^-5^ for genome wide association data.

We then examined associations of the same variants in ∼1 million human DNA samples spanning 23 diagnosed neuropsychiatric diseases and mental health traits using the same analytic approaches. For *F508del*-CFTR, we used GWAS summary statistics from 21 studies. When *F508del-CFTR* was not included in the study, we used the closely linked rs113827944 variant (r^2^ = 0.7 in Europeans) as a proxy. For CFTR premature termination variants, we used burden tests from whole-exome sequencing for 3 neuropsychiatric disease available through public browsers: autism spectrum disorder (ASC(21)), bipolar disorder (BipEx (22) and schizophrenia (SCHEMA (23)). In addition, we used data from Tian et al.(24) on depression and we curated a set of 12 diagnosed neuropsychiatric or mental health conditions with large number of cases from Genebass and All by All. The pre-specified threshold for genome-wide association is 10^-8^; based on 46 hypothesis tests (*F508del* and protein-truncating variants each for 23 neuropsychiatric diseases) the threshold for suggestive association is p <10^3^.

## Results

### Receptor binding studies at clinically relevant free concentrations did not identify any relevant off-target activity for ELX, VNZ, TEZ, IVA, or their major metabolites

No clinically relevant off-target binding activity was observed for ELX, VNZ, TEZ, IVA, or their major circulating metabolites across more than 65 neuropsychiatric-related receptors, ion channels, and transporters (**Table 1**). The clinically relevant *in vivo* concentration used is that of the free (unbound) concentration that interacts with the target, since protein-bound small molecules are not pharmacologically active.(15) The use of the clinically relevant free concentration (rather than total concentration) is particularly relevant to CFTRm because of the unusually high human plasma protein binding of ELX (99.3%), VNZ (99.99%), TEZ (99.1%), and IVA (99.95%). The high protein binding of CFTRm means the clinically relevant <u>free</u> concentration of each CFTRm is a small fraction of the total concentration. *In vitro* assays performed using total concentration in the absence of serum protein result in much higher free concentrations than are clinically relevant in the presence of total plasma protein.(25) Table 1 illustrates the large margin between the free concentration *in vitro* and the free concentration in the plasma at the therapeutically relevant C_max_ for ELX, VNZ, TEZ, and IVA.

**Table 1.**
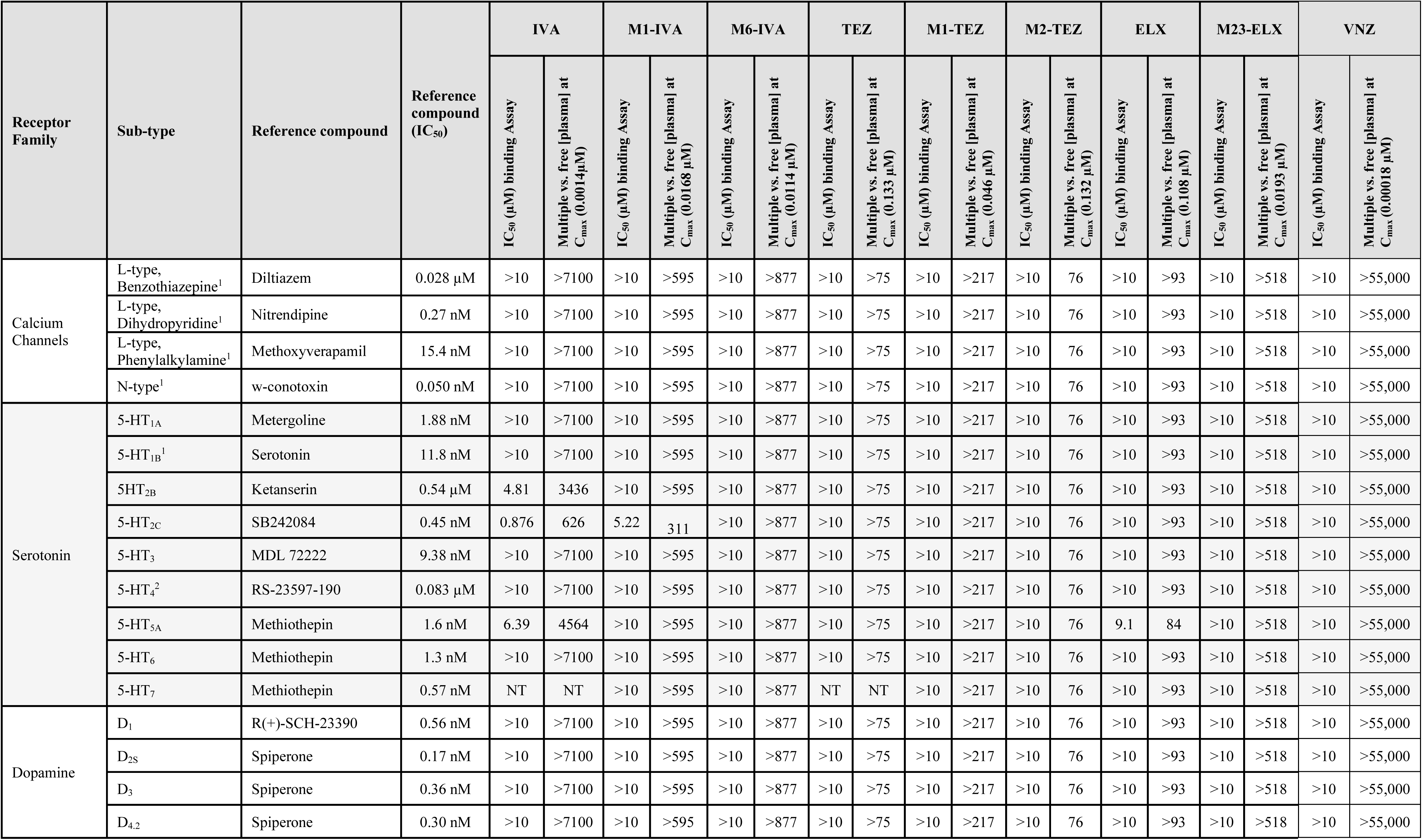
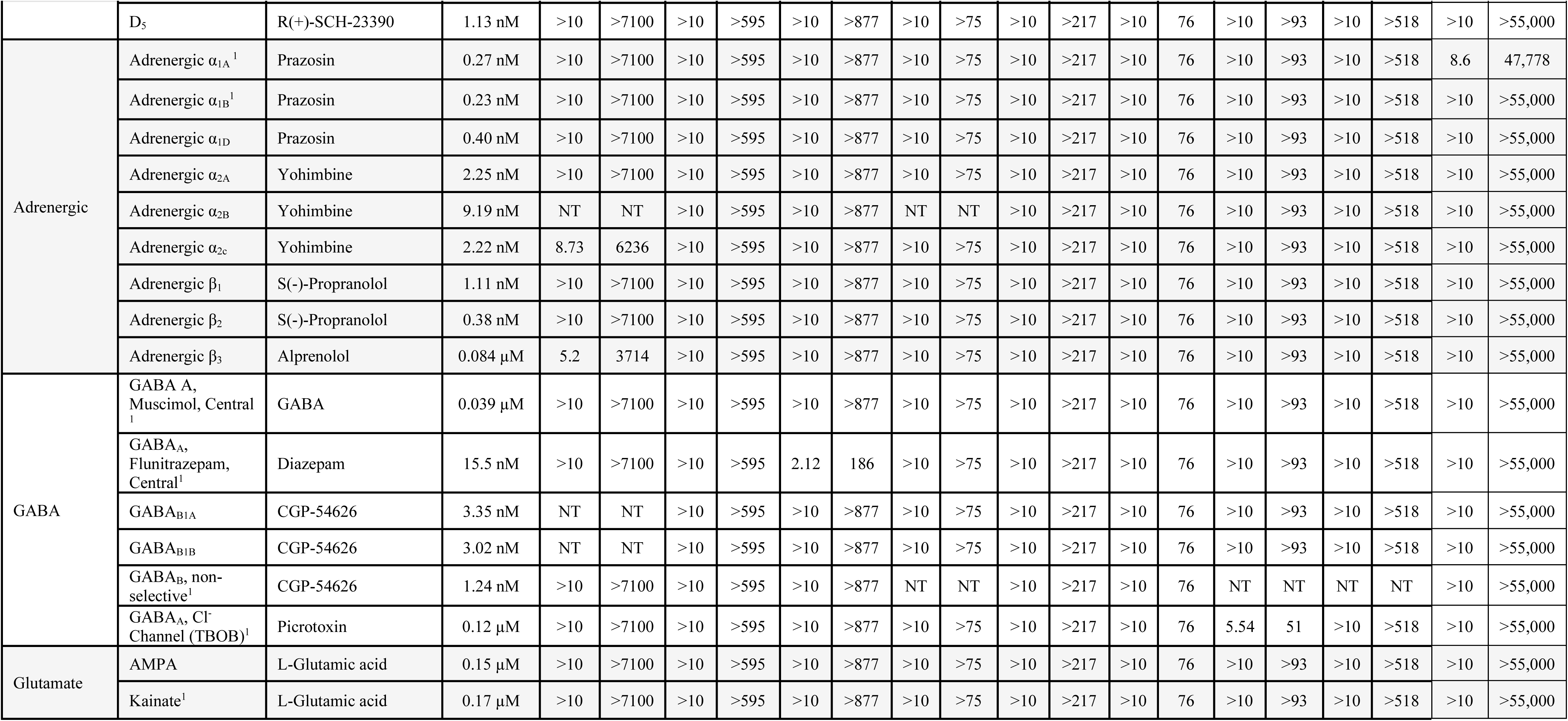
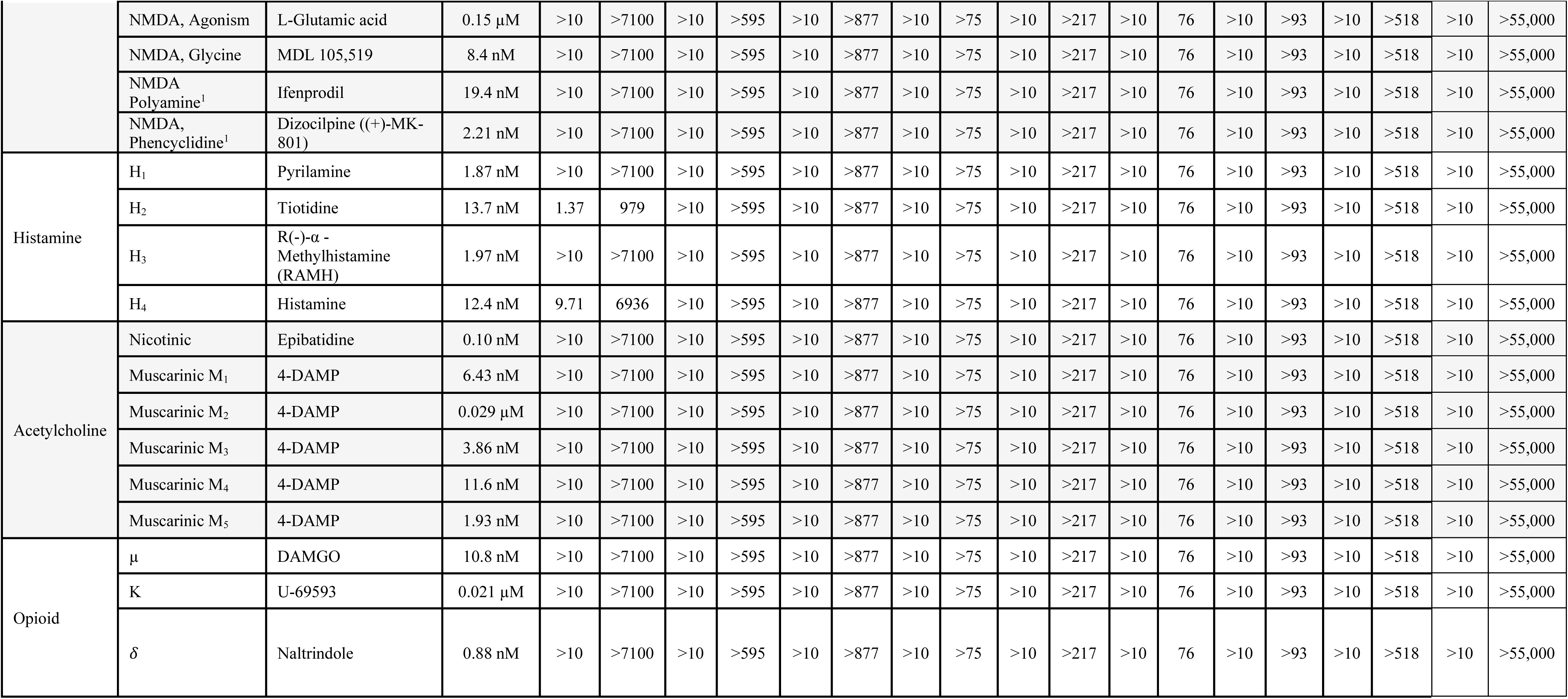
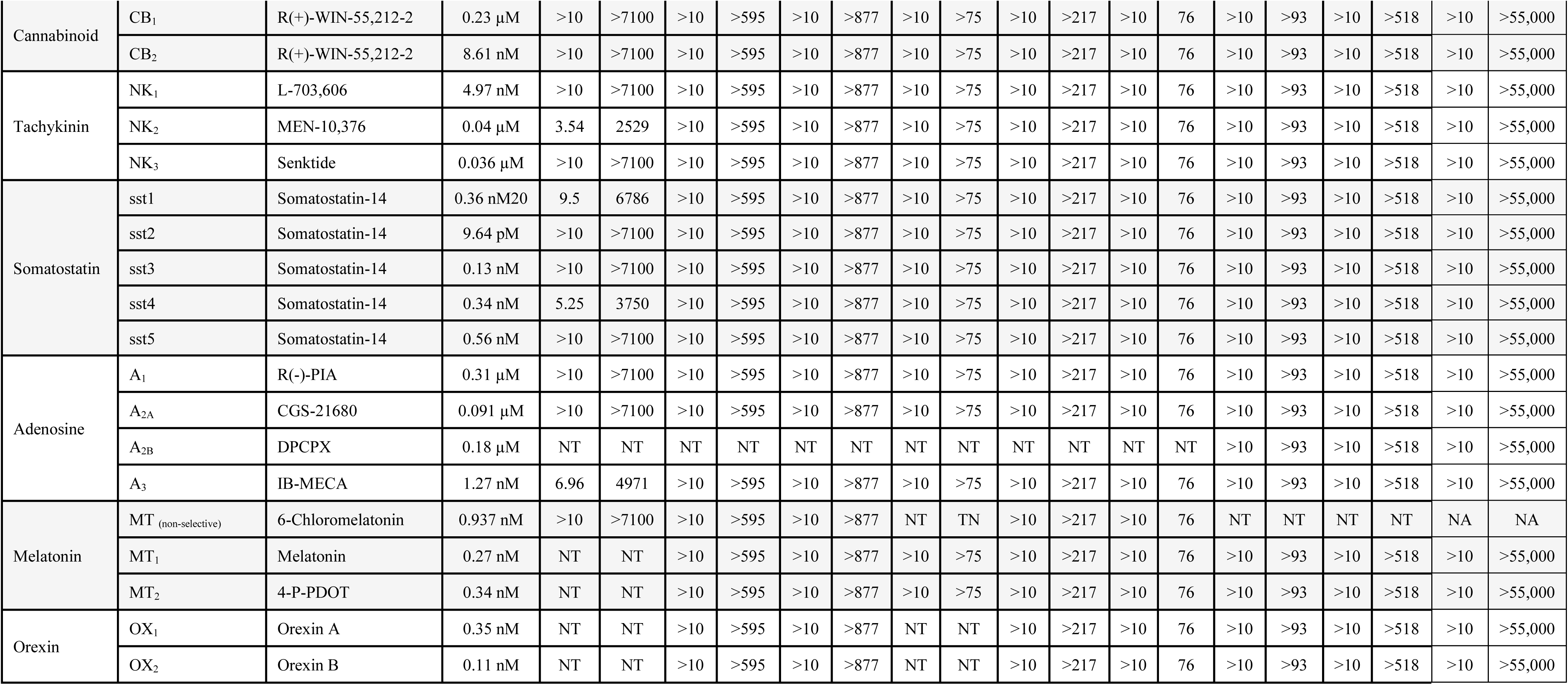
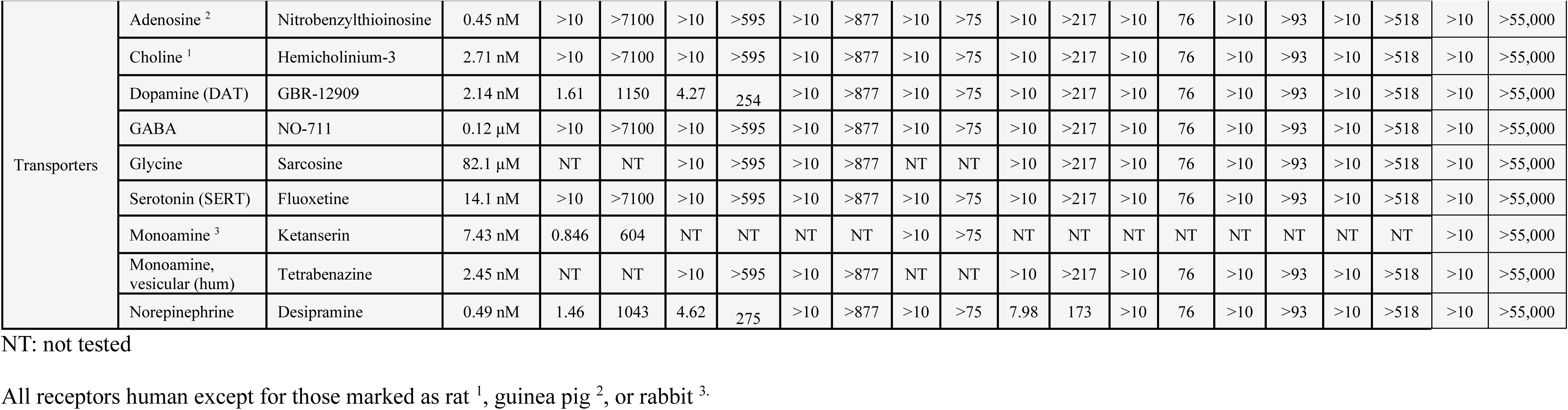
Selectivity of ELX, VNZ, TEZ, and IVA against major receptor and transporter families implicated in mental health disorders.

Given the hypothesis that CFTRm may have an effect on the brain, we evaluated the relative concentrations for CFTRm in central nervous system (CNS) as compared to plasma using rat ^14^C radiolabel tissue distribution studies. These studies showed that at C_max_ the brain concentrations of ELX, VNZ, TEZ and IVA were approximately 23%, 15%, 39%, and 3% of the concentration measured in plasma, respectively (**Table 2**). This potentially results in a larger margin between any potential off-target activity observed *in vitro* and clinically relevant free concentration in the CNS.

**Table 2.**
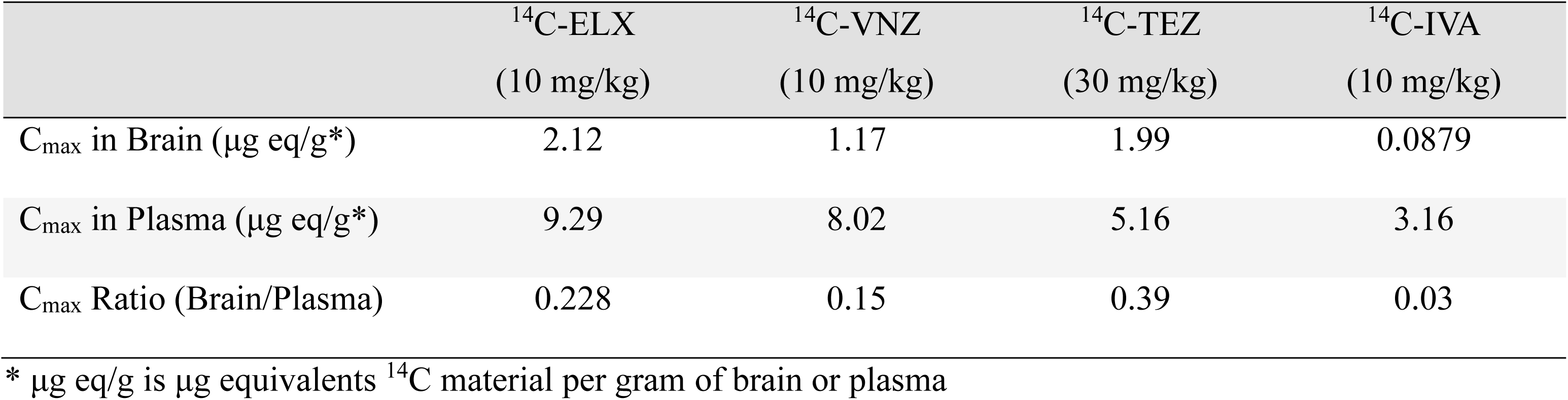
Tissue distribution of ^14^C-ELX, ^14^C-TEZ and ^14^C-IVA at the C_max_ following oral administration to Male Sprague Dawley Rats.

### Neurobehavioral studies in rats did not identify central nervous system behavioral changes following oral administration of ELX, VNZ, TEZ, or IVA

No neurobehavioral changes were observed in rats treated with ELX, VNZ, TEZ, or IVA across all dose groups and timepoints (**Table 3**). In the Irwin test, rats administered IVA showed no deviation from normal across all evaluated parameters at any dose or post-dose timepoint. To confirm assay sensitivity, chlorpromazine-treated rats served as a positive control and exhibited the expected signs of central nervous system depressant activity, including decreased body tone, decreased touch response, ptosis, and abnormal gait. Similarly, rats treated with ELX, VNZ, and TEZ demonstrated normal results across all FOB endpoints (activity/arousal, autonomic, neuromuscular, sensorimotor, and physiological) and were comparable to controls. No mortality or adverse clinical observations were reported for any CFTRm group. Collectively, these findings indicate that neither IVA (Irwin test) nor TEZ, ELX or VNZ (FOB) induced neurobehavioral effects in rats.

**Table 3.**
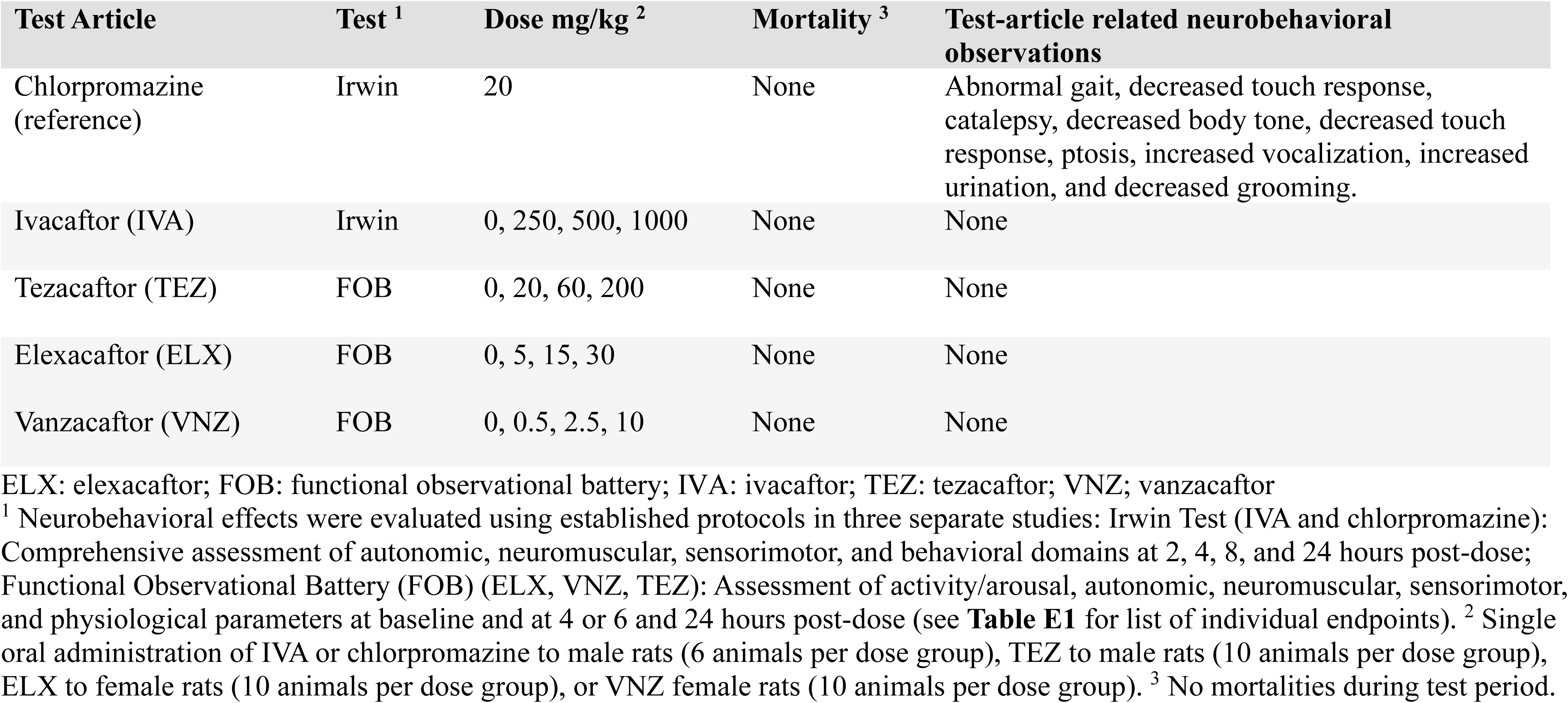
Clinical and FOB observations following oral gavage administration of Chlorpromazine, IVA, TEZ, ELX, or VNZ in female or male rats.

### Clinical trial and real-world data do not identify an association between CFTRm modulators and neuropsychiatric symptoms

To evaluate whether CFTRm affect neuropsychiatric symptoms, we analyzed data from clinical trials, real-world studies, and a literature review.

#### Clinical trial experience

Analysis of clinical trials with nearly 6,000 participants and over 12,000 patient-years (PY) of exposure showed no evidence linking CFTRm treatment (ELX/TEZ/IVA, TEZ/IVA, lumacaftor/IVA, and IVA) to neuropsychiatric events. In Phase 3 trials and their open-label extensions, rates of neuropsychiatric symptoms were low and similar between treatment and placebo groups (**Table 4**), and the nature of the neuropsychiatric events was similar across treatment groups (**Table E2**). Across 60 Phase 2 and 3 trials, the incidence and nature of the neuropsychiatric events were comparable between treated and placebo groups (**Table E3**). Notably, the rate of neuropsychiatric events in the treatment group was lower (6.9 events/100 PY) for CFTRm than in the pooled placebo group (9.6 events/100 PY). Serious neuropsychiatric events and those leading to treatment discontinuation were infrequent and occurred at similar rates between the treated and placebo groups.

**Table 4.** Exposure-Adjusted Summary of Relevant Psychiatric AEs Among CFTR Modulator Regimens: Pivotal Placebo-Controlled Phase 3 and Open-Label Extension Studies in pwCF.

| Events/<br>100PY | ELX/TEZ/IVA |  |  | TEZ/IVA |  |  | LUM/IVA |  |  | IVA |  |  |
| --- | --- | --- | --- | --- | --- | --- | --- | --- | --- | --- | --- | --- |
|  | 24w,<br>PBO-controlled |  | 4y, OLE | Pooled 24w,<br>PBO-controlled |  | 2y, OLE | Pooled 24w,<br>PBO-controlled |  | 2y, OLE | Pooled 48-wk,<br>PBO-controlled |  | 2y, OLE |
|  | PBO<br>N=201 | ETI<br>N=202 | ETI<br>N=506 | PBO<br>N = 505 | T/I<br>N=496 | T/I<br>N=1042 | PBO<br>N=370 | L/I<br>N=738 | L/I<br>N=1029 | PBO<br>N=104 | IVA<br>N=109 | IVA<br>N=192 |
| Relevant Psychiatric Events | 8.01 | 8.98 | 8.27 | 10.94 | 6.48 | 7.51 | 16.47 | 8.74 | 6.32 | 4.13 | 5.57 | 7.17 |
| Relevant Psychiatric Serious Events <sup>a</sup> | 2.00 | 0 | 0.38 | 0 | 0 | 0.72 | 1.10 | 0 | 0.41 | 1.03 | 0 | 0.80 |
| Relevant Psychiatric Events leading to Discontinuation | 0 | 0 | 0.05 | 0 | 0 | 0.13 | 0 | 0 | 0 | 1.03 | 0 | 0.27 |

AE: adverse event; CF: cystic fibrosis; CFTR: CF transmembrane conductance regulator protein; ETI: triple combination therapy with elexacaftor, tezacaftor, and IVA; IVA: ivacaftor; L/I: lumacaftor in combination with IVA; N: total sample size; OLE: open-label extension study; PBO: placebo; pwCF: people with CF; PY: person-year; SAE: serious AE; T/I: tezacaftor in combination with IVA; w: week. Events/100PY: number of events per 100 PY = number of events/total duration of safety analysis period in 100PY.
Notes: When summarizing number of events, a participant with multiple events within a category is counted multiple times in that category.
<sup>a</sup> Relevant psychiatric preferred terms comprise abnormal behavior, anxiety, completed suicide, depression, depressed mood, depression suicidal, emotional disorder, insomnia, mental disorder, mood altered, sleep disorder, suicide attempt, suicidal behavior, suicidal ideation, and suicide threat. Note that there were no reported events of abnormal behavior, completed suicide, mood altered, suicidal behavior and suicide threat in participants treated with CFTR modulators or placebo.

#### Post-authorization safety studies

A registry-based safety study sponsored by Vertex found that observed temporal patterns of depression or anxiety diagnoses prevalence did not change following the initiation of ELX/TEZ/IVA (**Figure 1**).

#### Literature search for CFTRm and depression- or anxiety-related events

A systematic literature review (SLR) identified 21 real world data publications from 8 countries (>1,300 participants) that systematically assessed depression (12 studies), anxiety (11 studies), sleep quality/disturbances (6 studies), and other topics (behavioral issues, emotional quality of life: 12 studies) using instruments such as the PHQ-9, GAD-7, and others (**Figure E1**). Key results are summarized in **Table E4**.

No study identified significant worsening of mental health outcomes at the population level following ELX/TEZ/IVA initiation (**Table E4**). Of 12 studies evaluating depression scores following initiation of ELX/TEZ/IVA treatment, 8 studies reported no change, 3 reported improvements, and one reported a mixed result depending on the instrument used. Among 11 studies evaluating anxiety, 9 found no change and 2 reported improvements. Sleep quality was assessed in 6 studies, with 3 showing no change and 2 showing improvement; an additional study using polysomnography found either no change or improvement across sleep indices. Emotional quality of life and behavioral outcomes also showed no change or improvement following CFTRm ELX/TEZ/IVA initiation. One of these studies evaluated behavioral patterns in children using the PSC-17 instrument and showed no change following ELX/TEZ/IVA initiation.

### Human genetic studies of CFTR functional variants did not identify any association across a broad spectrum of neuropsychiatric diseases in approximately 1 million individuals

We performed human genetic analysis of functional *CFTR* variants to address the hypothesis that CFTR expression and function in the CNS could be related to neuropsychiatric traits. This hypothesis could potentially explain the lack of findings in clinical trial data due to the relatively short duration of clinical trials and/or the lack of well-defined diagnostic criteria for neuropsychological traits used in CF clinical studies.

To evaluate this possibility we used human genetic data to test whether functional variants in *CFTR* with lifetime exposure have any association with large international cohorts diagnosed with a variety of neuropsychiatric diseases.

Specifically, we analyzed genetic data from 24 neuropsychiatric studies in approximately 1 million individuals as well as 3 population wide biobanks to search for any phenotypic associations of *CFTR* functional variants (**Figure 2; Tables E5 and E6**). The human genetic analyses utilized two distinct and complementary sets of *CFTR* functional variants: 1) genotype of the common functional variant *F508del-CFTR*, and 2) pooled Class I premature termination variants in *CFTR* identified by exome sequencing data. This analysis is based on the principle of Mendelian Randomization (26), where inherited genetic variants affecting the function of a gene provide an unconfounded assessment of the relationship between the gene’s function and a clinical phenotype of interest.

**Figure 2.**
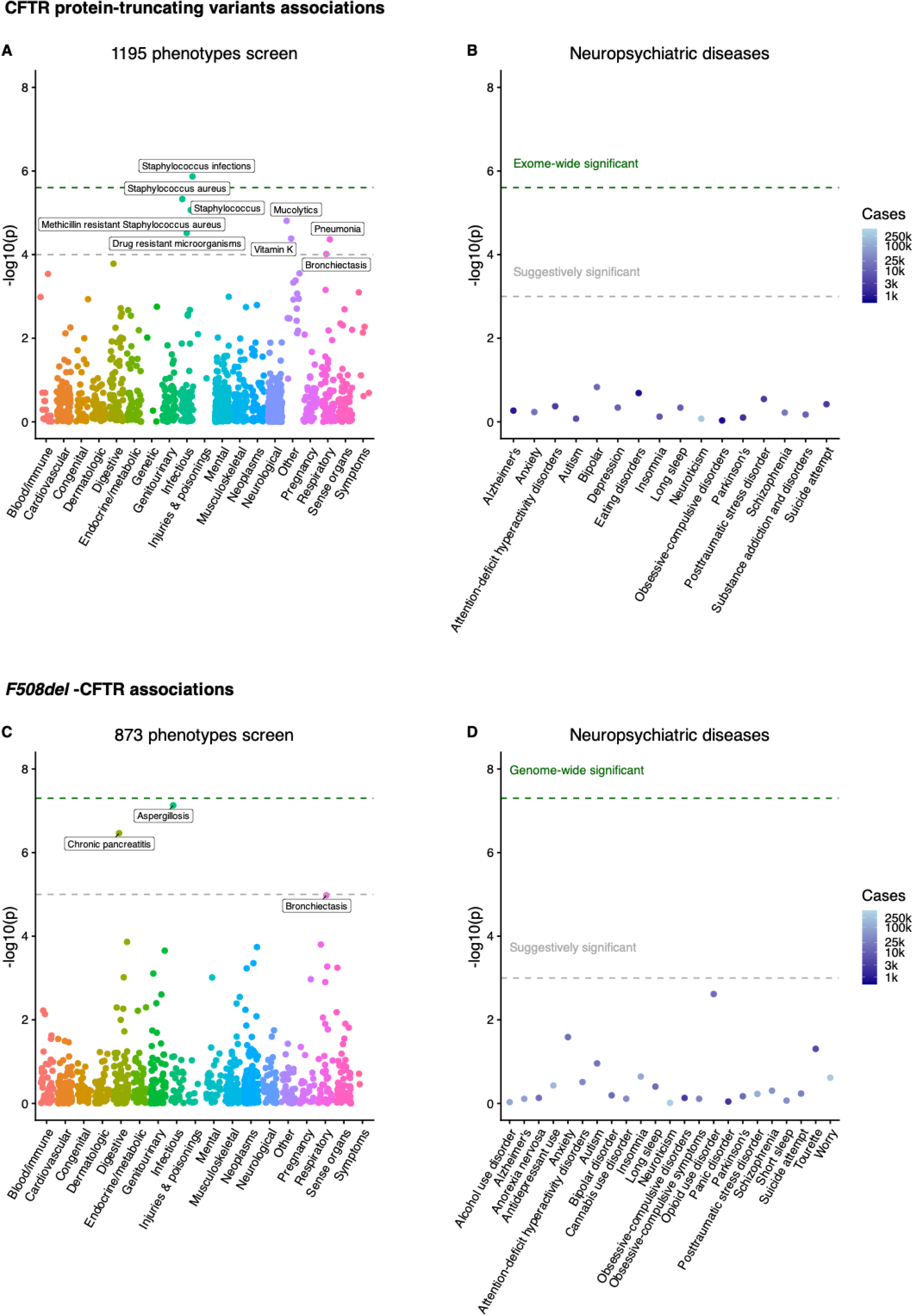
Lack of human genetic association between *CFTR* variants and a broad spectrum of neuropsychiatric conditions in ∼1 million individuals. A. Results of gene burden test studies for class I *CFTR* protein-truncating variants across an unbiased set of 1,195 phenotypes from two biobanks comprising 644k samples in total. Each dot represents a phenotype and is placed on the x-axis and colored according to its category. The y-axis shows the p-value of the association, the grey and green dashed lines indicate the suggestive (p < 1e-4) and exome-wide (p < 2.5e-6) significance thresholds. Phenotypes that are associated with p < 1e-4 are labelled. B. Results from class I *CFTR* protein-truncating variants burden tests in 16 neuropsychiatric disorders and mental health traits from 6 studies with up to 18,000 cases. The y-axis shows the p-value of the association, the grey and green dashed lines indicate the suggestive (p < 1e-3) and exome-wide (p < 2.5e-6) significance thresholds. The color indicates the number of cases included in the study. C. GWAS association results between *F508del* and an unbiased set of 873 phenotypes in a biobank meta-analysis comprising over 1M individuals. Each dot represents a phenotype and is placed on the x-axis and colored according to its category. The y-axis shows the p-value of the association, the grey and green dashed lines indicate the suggestive (p < 1e-5) and genome-wide (p < 5e-8) significance thresholds. Phenotypes that are associated with p < 1.1e-5 are labelled. D. GWAS association results between *F508del* and 23 neuropsychiatric disorders and mental traits from 21 studies with up to 192k cases. The y-axis shows the p-value of the association, the grey and green dashed lines indicate the suggestive (p < 1e-3) and exome-wide (p < 5e-8) significance thresholds. The color indicates the number of cases included in the study.

As a positive control we first used phenome-wide scans in public biobanks data to evaluate whether the *CFTR* functional variants were associated with any human traits (See methods, **Figure 2**, **Table E5**). For *F508del-CFTR* we analyzed a meta-analysis of two large public biobanks including approximately 1 million individuals and 873 phenotypes. For the *CFTR* premature termination variants analysis, we used exome sequencing data from two biobanks totaling 645,000 individuals and 1,195 phenotypes.

These analyses in two large public biobanks showed that 1 *F508del* was associated with an increased risk of aspergillosis (beta = 1.05, p = 7.41e-8) and chronic pancreatitis (beta = 0.588, p = 3.43e-7) and that pooled Class I *CFTR* premature termination variants were associated with Staphylococcus infections (beta = 4.83, p = 1.35e-6), mucolytics (beta = 4.32, p = 1.55e-5), drug resistant microorganisms (beta = 4.17, p = 3.04e-5), vitamin K deficiency (beta = 4.10, p = 4.14e-5), bronchiectasis (beta = 3.90, p = 9.64e-5) and pneumonia (beta = 4.09, p = 4.33e-5). The fact that unbiased phenome scanning of CF-causing *CFTR* variants identified only established CF-related phenotypes supports the sensitivity of the analysis performed and the validity of this approach.

We next performed the same association methods (*F508del* and pooled *CFTR* premature termination variants) across 20 genome-wide association and 4 exome sequencing studies of diagnosed patients and controls for 23 neuropsychiatric or mental health diagnoses (**Figure 2, Table E6**). These studies were performed by consortia that have successfully discovered genetic variants associated with each neuropsychiatric condition and in total include ∼1 million cases of one or more psychiatric diseases. The different studies span sample sizes ranging from 10,000 to >1 million participants, with case counts ranging from 660 to 191,800, diagnosed patients. For the pooled *CFTR* premature termination variants, we also used 12 curated phenotypes from the two biobanks as described above because sequencing data from disease cohorts were not available.

Results of the analysis of *F508del-CFTR* and of Class 1 *CFTR* variants identified no associations that met the pre-specified statistical threshold for any of the neuropsychiatric or mental health traits.

Finally, we used public data from the Genotype-Tissue Expression (GTEX; https://www.gtexportal.org/home/snp/rs10255092, consulted Sep. 19^th^, 2025) to identify a common non-CF-causing variant (rs10255092) upstream of *CFTR* that is associated with expression of *CFTR* mRNA in the CNS. Association studies of this variant that modifies CFTR expression in the CNS showed no association with any of the 23 neuropsychiatric or mental health traits investigated (**Table E6**).

These human genetic studies identified associations of *CFTR* variants to traits known to be associated with CFTR function and CNS expression, but did not identify any association of *CFTR* variants with neuropsychiatric traits.

## Discussion

A central question of current care for pwCF is whether there is a relationship between neuropsychiatric symptoms and either off-target or on-target effects of CFTRm therapy. To address this question, we conducted a comprehensive assessment that included *in vitro* receptor binding studies, *in vivo* animal behavioral pharmacology studies, clinical trial and real-world data, and human genetic data from ∼1 million individuals diagnosed with one or more neuropsychiatric diseases. These studies did not find any *in vitro* evidence of off-target effects at clinically relevant concentrations, central nervous system behavioral changes in rats, effects of treatment with ELX/TEZ/IVA or VNZ/TEZ/D-IVA in clinical trials or real-world settings, or genetic evidence linking CFTR functional mutations to a broad spectrum of mental health disorders. The totality of the data fails to support the hypothesis that CFTRm directly exert an on-target or off-target effect on neuropsychiatric traits.

To interpret the *in vitro* findings it is necessary to address the free drug concentrations in the plasma, not the total concentrations.(27) For example, at the C_max_ for 150 mg bid IVA when administered in combination with ELX and TEZ, the total and free plasma concentrations of IVA are 3.057 µM and ∼0.0014 µM, respectively. In the receptor binding assay (performed without serum protein), the IC_50_ for IVA against a broad spectrum of neuropsychiatric-related targets ranged from 0.85 µM to >10 µM. This indicated that there is a ∼600-to->7100-fold margin between the *in vitro* IC_50_ and the clinically relevant free plasma concentration for IVA. In published work the effects of ELX on large-conductance calcium channels were tested at 10 µM in the absence of serum protein(12) — a more than ∼100-fold increase above the clinically relevant free concentration *in vivo*. Similarly, Scheider et al.(11) reported binding of IVA, M1-IVA and M6-IVA to 5-HT_2_ receptors with a K_i_ = 0.866 µM, which is ∼600 fold higher than the clinically relevant free concentration. The high protein binding of ELX, VNZ, TEZ, and IVA, and the low free concentrations of each, mean that the *in vitro* activity previously reported in assays lacking serum protein results in a very large margin compared to the clinically relevant free concentrations. Free concentrations in the CNS may be further reduced by the low predicted brain exposures observed in rat tissue distribution studies of ELX, VNZ, TEZ, and IVA.

To assess the potential biological role of CFTR in neuropsychiatric behavior, we examined associations between *CFTR* variants and a broad spectrum of phenotypes, including 23 neuropsychiatric or mental health diagnoses, in ∼ 1 million individuals. Consistent with prior studies, *CFTR* variants were associated with pancreatitis, aspergillosis, and bronchiectasis.(28) In contrast, no associations were identified between *CFTR* variants and a broad spectrum of mental health diagnoses, including depression, anxiety, and cognitive impairment. Taken together, these genetic data do not support the hypothesis that alteration of CFTR function by on-target activity of CFTRm would be expected to influence neuropsychiatric symptoms.

Given the lack of evidence reported in this study for off-target or on-target effect of CFTRm on neuropsychiatric disease, it is appropriate to consider alternative hypotheses for the reported neuropsychiatric symptoms in some individuals with CF. One possibility is a secondary effect of changes in physical health and life expectancy on personal identity and needs. Transformative therapies in other serious diseases, such as antiretroviral treatment for HIV, organ transplantation, and cancer have significantly improved the quality and duration of life, but they may also trigger new or worsening neuropsychiatric symptoms. (29–31) A similar phenomenon could contribute to some of what is occurring with CFTRm. (32, 33).

This study has important limitations. While clinical trial and registry data encompass large populations and extensive patient-years, detailed neuropsychiatric assessments were not systematically included in multi-center trials, making it challenging to detect rare or subtle effects. Smaller studies using validated instruments have not identified significant associations, but these are often unblinded and uncontrolled. Importantly, the absence of direct symptom queries in large datasets may reduce ascertainment bias, yet it also limits the ability to capture nuanced patient experiences. As such, while population-level analyses do not support a direct link between CFTRm and neuropsychiatric symptoms, and anecdotal cases are often confounded by incomplete data and the fluctuating nature of neuropsychiatric symptoms, rare individual cases cannot be fully excluded. Additionally, not every potential neuropsychiatric target or disorder was examined in these analyses, leaving open the possibility that effects on unstudied conditions may not have been detected. Prior analyses have examined depression-related events, and the analyses presented here include a broader array of neuropsychiatric symptoms.(8) The genetic studies of CFTR functional variants in large diagnosed cohorts of patients with neuropsychiatric diseases were intended to provide an alternative approach to evaluating the role of CFTR that does not share the limitations of the clinical and real world evidence.

In conclusion, *in vitro* and preclinical *in vivo* studies, clinical trial and real-world results, and human genetic data did not identify evidence of off-target binding of ELX, VNZ, TEZ or IVA at clinically relevant concentrations, behavioral symptoms in animals, mental health symptoms in clinical trials, or association of CFTR functional variants with neuropsychiatric traits.

## Data Availability

All data produced in the present study are available upon reasonable request to the authors

## Acknowledgments

The authors thank the patients and their families for participating in this trial and the study investigators and coordinators for their contributions to the study. Medical writing and editorial support were provided by Crystal Kellas, Ph.D., and graphical assistance was provided by Patrick Coughlin, under the guidance of the authors. We also thank John Pettersen and Jeffrey Wiley for their review of the manuscript. FVG, DA, PS, JK, LF, NA, WC are employees of Vertex Pharmaceuticals Incorporated and may own stock or stock options in the company.

---

**Figure E1.**
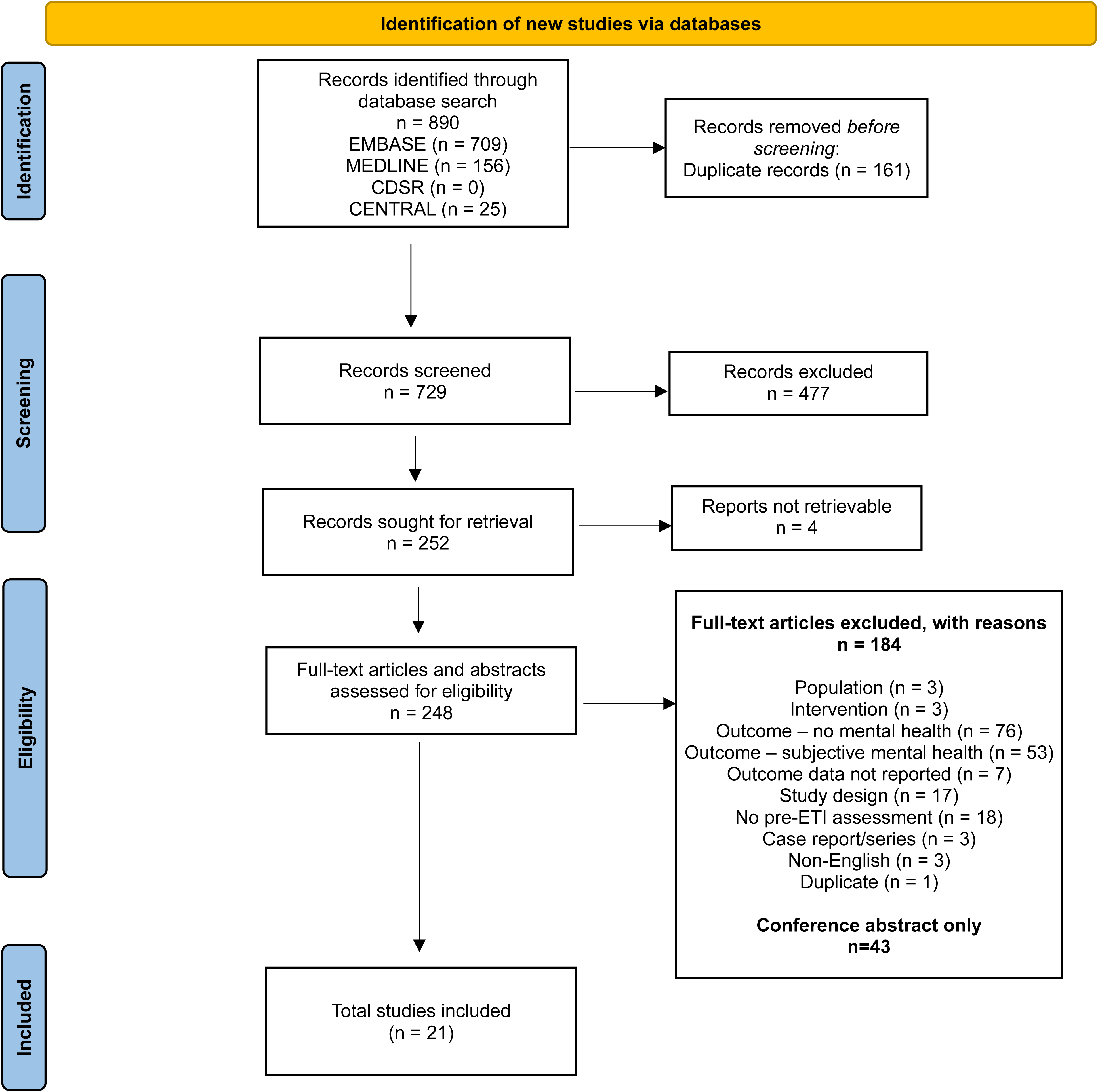
PRISMA flow diagram.

