## Supplemental Appendix for "Evaluation of a Potential Relationship between CFTR Modulators, CFTR function and Neuropsychiatric Symptoms in People with CF"

### Supplementary Methods

#### Rat neurobehavioral studies

*Animal studies and dosing*

Sprague Dawley rats (male or female, depending on compound) at least 6 weeks of age at study start were acclimated and randomly assigned to treatment groups. Each CFTR modulator—elexacaftor (ELX), vanzacaftor (VNZ), tezacaftor (TEZ), or ivacaftor (IVA)—was administered as a single oral gavage at three dose levels, with vehicle controls included in all studies. For the IVA study, a positive control group received chlorpromazine (20 mg/kg) to confirm assay sensitivity. Animal behavioral studies were conducted by Aptuit, Inc. Kansas City Missouri (Irwin test: IVA) or MPI Research, Inc. Mattawan, Michigan (FOB: TEZ and ELX) in accordance with the United States Food and Drug Administration Good Laboratory Practice Standards.

*Behavioral assessments*

Neurobehavioral effects were evaluated using established protocols: 1) Irwin Test (IVA): Comprehensive assessment of autonomic, neuromuscular, sensorimotor, and behavioral domains at 2, 4, 8, and 24 hours post-dose; Functional Observational Battery (FOB) (ELX, VNZ, TEZ): Assessment of activity/arousal, autonomic, neuromuscular, sensorimotor, and physiological parameters at baseline and at 4 or 6 and 24 hours post-dose (**Table E1**). Body weight and temperature were monitored throughout. All animals were observed for morbidity, mortality, and general health at least twice daily.

*Formulation and Data Analysis*

Test articles were suspended in methylcellulose-based vehicle and administered within hours of preparation. Dosing formulations were analyzed for concentration and homogeneity using validated HPLC-UV methods; all met acceptance criteria.

Data were compared between treated and control groups to identify neurobehavioral changes. For FOB endpoints (ELX, VNZ, TEZ), statistical comparisons between treatment and vehicle treated groups included Group Pair-wise Comparisons (continuous endpoints) or Cochran Mantel Haenszel Test (Categorical Endpoints). For the Irwin test (IVA, Chlorpromazine), only deviations from normal were recorded.

*Study Termination*

At study completion, animals were euthanized by carbon dioxide inhalation followed by a secondary method to confirm death. No postmortem evaluations were scheduled unless required for unexpected deaths.

**Determination of the free concentration at the $C_{max}$ for ELX, VNZ, TEZ, and IVA**

The free fractions of ELX, VNZ, TEZ, and IVA in human plasma were determined using equilibrium dialysis method. The 3 mM stock solutions of ELX, VNZ, TEZ, and IVA were prepared from compound powder using DMSO. Diluted plasma was prepared by adding an appropriate amount of blank plasma to the dialysis phosphate buffer. Into separate vials for each compound were spiked into diluted plasma at a final concentration of 5 µM (1% organic). The control compound, warfarin, was spiked into the undiluted plasma at 5 µM. The spiked diluted or undiluted plasma was loaded into the plasma compartment (red side) of the Rapid Equilibrium Dialysis (RED) device (200 µL; n = 4 per condition). Phosphate buffer was loaded into the buffer compartment (white side) of the RED device (350 µL; n = 4 per condition). The RED device was sealed with breathable sealing tape and then an adhesive sealing tape from ThermoFisher Scientific (Waltham, MA). The device was agitated gently on a shaking platform at 150 rpm in a $CO_2$ incubator (5% $CO_2$) for 18 hours at 37°C with saturating humidity. After 18-hour incubation, 20 µL from the plasma side and 20 µL from the buffer side of each RED well were added into an equal amount of the opposite blank matrix. Samples were mixed with 480 µL of Internal Standard solution in acetonitrile to precipitate proteins (i.e., 20 µL of sample plasma + 20 µL of blank sodium phosphate buffer; 20 µL of sample buffer + 20 µL of blank diluted plasma). Samples were vortexed and centrifuged at 12000 rpm for 11 minutes. The supernatants were injected for LC-MS/MS analysis. Compounds bound to plasma proteins do not cross the dialysis membrane. The fraction bound can thus be calculated from the difference between the compound concentrations in the plasma (donor) and buffer (receiver) samples.

**Rat brain distribution studies**

The tissue distribution of $^{14}$C-IVA was studied in male Sprague-Dawley rats following oral administration at 10 mg/kg with the dose formulated polyethylene glycol (PEG) 400 with a dose volume of 5 mL/kg. The targeted radioactive dose of $^{14}$C-IVA was 100 µCi/kg. Tissue concentrations were determined at 1, 4, 24, 48,72, and 168 hours post dosing.

The tissue distribution of $^{14}$C-TEZ was studied in male Sprague-Dawley rats following oral administration at 30 mg/kg with the dose formulated in 37.5% PEG 400/12.5% Vitamin E-TPGS in water with a dose volume of 5 mL/kg. The targeted radioactive dose of $^{14}$C-TEZ was 100 µCi/kg. Tissue concentrations were determined at 1, 4, 24, 48, 72, and 168 hours post dosing.

The tissue distribution of $^{14}$C-ELX was studied in male Sprague Dawley rats following oral administration at 10 mg/kg with the dose formulated in 5% NMP/30% PEG 400/10% TPGS/5%

PVP VA in water with a dose volume of 10 mL/kg. The targeted radioactive dose of $^{14}$C-VX-445 was 100 µCi/kg. Tissue concentrations were determined at 1, 4, 24 and 168 hours post dosing.

**Systematic Review of Published Literature**

The SLR search included peer-reviewed publications published from 2019 (first ELX/TEZ/IVA approval in the US) through May 7, 2025 (date of search) with the following key criteria: includes people with a diagnosis of CF treated with ELX/TEZ/IVA, reports on mental health outcomes measured using validated instruments or objective pre-defined patient- or caregiver-reported measures, and has data available prior to and after ELX/TEZ/IVA initiation (to assess post-treatment changes). VNZ/TEZ/D-IVA was excluded as it was recently approved and similar studies have not been conducted.

Searches for this SLR were conducted across multiple databases with the Ovid® search interface (Embase, Ovid MEDLINE®, and the Cochrane Central Register of Controlled Trials) using a combination of controlled vocabulary and keywords by a medical information specialist and peer reviewed by another senior medical information specialist before execution (**Figure E1**). The initial search was not restricted by language. Study eligibility was assessed according to pre-specified Population, Intervention, Comparator, Outcome, and Study Design (PICOS) criteria (1) and was performed in duplicate by two independent reviewers. Data from included records were extracted using a standardized approach by a single reviewer. Data were then independently assessed for accuracy and completeness by a second reviewer. All conflicts were resolved through consensus or by a third independent reviewer.

**Clinical Trials**

Clinical trials referenced in this supplement were conducted in accordance with the Declaration of Helsinki, local applicable laws and regulations, and current Good Clinical Practice Guidelines as described by the International Council for Harmonisation.

**Figure E1. PRISMA flow diagram**

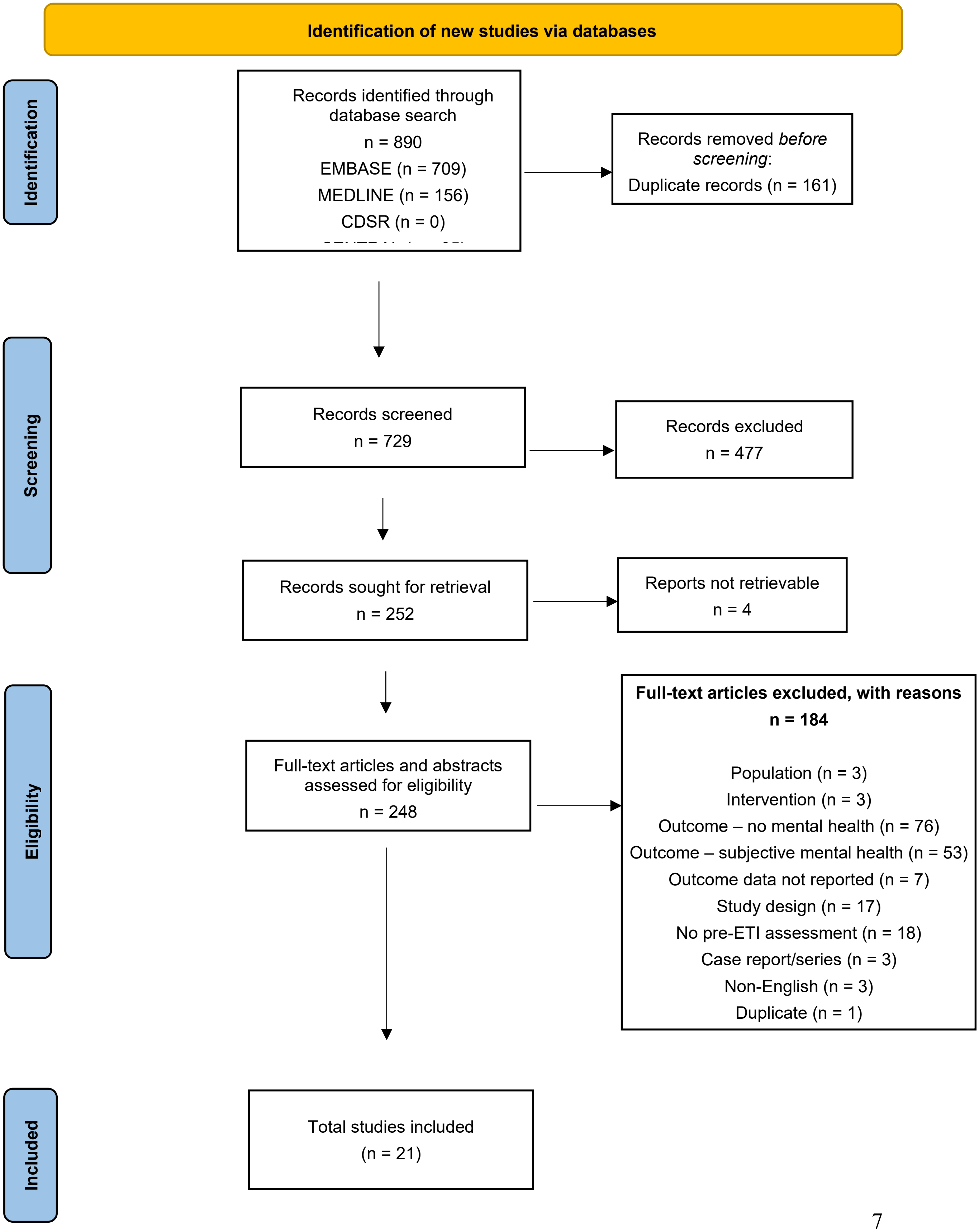

**Table E1. Functional neurobehavioral domains assessed for IVA (Irwin test) or TEZ, ELX, and VNZ (FOB tests)**

| Domain | Irwin Test Items | FOB Items |
|---|---|---|
| Mortality & General Health | Lethality, Cyanosis, Hypothermia | Body weight, Body temperature |
| Locomotor & Activity | Locomotor activity, Restlessness, Lethargy, Grooming, Dispersion in cage | Rearing, Posture, Arousal |
| Neuromuscular Function | Tremor, Twitches, Convulsions, Abnormal carriage/gait, Paralysis, Grip strength, Body tone, Catalepsy/passivity, Straub tail | Tremor, Convulsions, Forelimb/hindlimb grip strength, Hindlimb splay, Clonic/tonic movements, Gait, Stereotypy |
| Behavioral & Psychiatric-like Signs | Stereotypic behavior, Fighting, Aggressiveness, Fearfulness, Vocalization | Handling reactivity, Ease of removal, Vocalization, Bizarre behavior |
| Reflexes & Sensorimotor | Startle response/loss of righting reflex, Pinna reflex, Corneal reflex, Pain response | Thermal response, Approach/touch/click/tail pinch response, Pupil response, Righting reflex |
| Autonomic | Salivation, Lacrimation, Ptosis, Pupil diameter, Diarrhea, Increased urination, Cutaneous blood flow, Piloerection, Exophthalmia | Defecation, Urination, Lacrimation, Palpebral closure, Piloerection, Exophthalmos, Salivation, Pupil response |
| Physiological | Respiration, Cyanosis, Hypothermia, Body tone | Respiration, Body weight, Body temperature |
| Other | Writhing, Catalepsy/passivity, Cutaneous blood flow | |

ELX: elexacaftor; FOB: functional observed battery; IVA: ivacaftor; TEZ: tezacaftor; VNZ: vanzacaftor

**Table E2 Exposure-Adjusted Summary of Relevant Psychiatric AEs by Preferred Term Among CFTR Modulator Regimens: Pivotal Placebo-Controlled Phase 3 and Open-Label Extension Trials in pwCF**

| Events/ 100PY PT | ELX/TEZ/IVA | | | TEZ/IVA | | | LUM/IVA | | | IVA | | |
|---|---|---|---|---|---|---|---|---|---|---|---|---|
| | 24w, PBO-controlled | | 4y, OLE | Pooled 24w, PBO-controlledf | | 2y, OLE | Pooled 24w, PBO-controlled | | 2y, OLE | Pooled 48-wk, PBO-controlled | | 2y, OLE |
| | PBO N=201 | ETI N=202 | ETI N=506 | PBO N = 505 | T/I N=496 | T/I N=1042 | PBO N=370 | L/I N=738 | L/I N=1029 | PBO N=104 | IVA N=109 | IVA N=192 |
| Relevant Psychiatric Events[a] | 8.01 | 8.98 | 8.27 | 10.94 | 6.48 | 7.51 | 16.47 | 8.74 | 6.32 | 4.13 | 5.57 | 7.17 |
| Anxiety | 1.00 | 2.99 | 2.79 | 3.45 | 0.59 | 2.17 | 2.20 | 2.26 | 2.09 | 1.03 | 1.86 | 3.19 |
| Depression | 2.00 | 1.00 | 2.57 | 2.30 | 1.18 | 2.11 | 3.84 | 1.13 | 1.85 | 1.03 | 0 | 1.06 |
| Depressed mood | 0 | 1.00 | 0.11 | 0 | 0 | 0.20 | 0 | 0.85 | 0.35 | 0 | 0 | 0 |
| Depression suicidal | 0 | 0 | 0 | 0 | 0 | 0 | 0 | 0 | 0 | 0 | 0 | 0.27 |
| Emotional disorder | 0 | 0 | 0 | 0 | 0 | 0 | 0.55 | 0 | 0 | 0 | 0 | 0 |
| Insomnia | 1.00 | 2.99 | 2.14 | 4.03 | 3.54 | 2.70 | 8.23 | 3.38 | 1.80 | 1.03 | 2.78 | 1.59 |
| Mental disorder | 0 | 0 | 0.05 | 0 | 0 | 0 | 0 | 0.28 | 0 | 0 | 0 | 0 |
| Mood altered | 0 | 0 | 0 | 0 | 0 | 0 | 0 | 0.28 | 0 | 0 | 0 | 0 |
| Sleep disorder | 3.00 | 1.00 | 0.27 | 1.15 | 1.18 | 0.07 | 0.55 | 0.28 | 0.06 | 1.03 | 0.93 | 0.80 |
| Suicidal ideation | 1.00 | 0 | 0.16 | 0 | 0 | 0.07 | 0.55 | 0.28 | 0.12 | 0 | 0 | 0.27 |
| Suicide attempt | 0 | 0 | 0.16 | 0 | 0 | 0.20 | 0.55 | 0 | 0.06 | 0 | 0 | 0 |

| Events/ 100PY PT | ELX/TEZ/IVA 24w, PBO-controlled | | ELX/TEZ/IVA 4y, OLE | TEZ/IVA Pooled 24w, PBO-controlledf | | TEZ/IVA 2y, OLE | LUM/IVA Pooled 24w, PBO-controlled | | LUM/IVA 2y, OLE | IVA Pooled 48-wk, PBO-controlled | | IVA 2y, OLE |
|---|---|---|---|---|---|---|---|---|---|---|---|---|
| | PBO N=201 | ETI N=202 | ETI N=506 | PBO N = 505 | T/I N=496 | T/I N=1042 | PBO N=370 | L/I N=738 | L/I N=1029 | PBO N=104 | IVA N=109 | IVA N=192 |
| Relevant Psychiatric Serious Events | 2.00 | 0 | 0.38 | 0 | 0 | 0.72 | 1.10 | 0 | 0.41 | 1.03 | 0 | 0.80 |
| Anxiety | 0 | 0 | 0.16 | 0 | 0 | 0.33 | 0 | 0 | 0.12 | 1.03 | 0 | 0 |
| Depression | 1.00 | 0 | 0.05 | 0 | 0 | 0.13 | 0 | 0 | 0.17 | 0 | 0 | 0.27 |
| Depression suicidal | 0 | 0 | 0 | 0 | 0 | 0 | 0 | 0 | 0 | 0 | 0 | 0.27 |
| Mental disorder | 0 | 0 | 0.05 | 0 | 0 | 0 | 0 | 0 | 0 | 0 | 0 | 0 |
| Suicidal ideation | 1.00 | 0 | 0 | 0 | 0 | 0.07 | 0.55 | 0 | 0.06 | 0 | 0 | 0.27 |
| Suicide attempt | 0 | 0 | 0.11 | 0 | 0 | 0.20 | 0.55 | 0 | 0.06 | 0 | 0 | 0 |
| Relevant Psychiatric Events leading to Discontinuation | 0 | 0 | 0.05 | 0 | 0 | 0.13 | 0 | 0 | 0 | 1.03 | 0 | 0.27 |
| Anxiety | 0 | 0 | 0 | 0 | 0 | 0.07 | 0 | 0 | 0 | 1.03 | 0 | 0 |
| Depression | 0 | 0 | 0.05 | 0 | 0 | 0.07 | 0 | 0 | 0 | 0 | 0 | 0 |
| Depression suicidal | 0 | 0 | 0 | 0 | 0 | 0 | 0 | 0 | 0 | 0 | 0 | 0.27 |

| Events/ 100PY PT | ELX/TEZ/IVA | | | TEZ/IVA | | | LUM/IVA | | | IVA | | |
|---|---|---|---|---|---|---|---|---|---|---|---|---|
| | 24w, PBO-controlled | | 4y, OLE | Pooled 24w, PBO-controlledf | | 2y, OLE | Pooled 24w, PBO-controlled | | 2y, OLE | Pooled 48-wk, PBO-controlled | | 2y, OLE |
| | PBO N=201 | ETI N=202 | ETI N=506 | PBO N = 505 | T/I N=496 | T/I N=1042 | PBO N=370 | L/I N=738 | L/I N=1029 | PBO N=104 | IVA N=109 | IVA N=192 |

AE: adverse event; CF: cystic fibrosis; CFTR: CF transmembrane conductance regulator protein; ETI: elexcaftor/tezacaftor/ivacaftor, and IVA; IVA: ivacaftor; L/I: lumacaftor in combination with IVA; N: total sample size; OLE: open-label extension study; PBO: placebo; PY: person-year; SAE: serious AE; T/I: tezacaftor in combination with IVA; y: year(s). Events/100PY: number of events per 100 PY = number of events/total duration of safety analysis period in 100PY.

Notes: When summarizing number of events, a participant with multiple events within a category is counted multiple times in that category.

[a] Relevant psychiatric preferred terms comprise abnormal behavior, anxiety, completed suicide, depression, depressed mood, depression suicidal, emotional disorder, insomnia, mental disorder, mood altered, sleep disorder, suicide attempt, suicidal behavior, suicidal ideation, and suicide threat. Note that there were no reported events of abnormal behavior, completed suicide, mood altered, suicidal behavior and suicide threat in participants treated with CFTR modulators or placebo.

**Table E3 Characteristics and Demographics of Relevant Psychiatric Preferred Terms for IVA, LUM/IVA, TEZ/IVA, and ELX/TEZ/IVA from Ph2 and Ph3 Clinical Trial Experience in pwCF**

| Parameter | | Placebo (Pooled - all CFTRm) N= 1,887 n (%) | ELX/TEZ/IVA N= 2,032[c] n (%) | TEZ/IVA N= 1,849[c] n (%) | LUM/IVA N= 1,770[c] n (%) | IVA N= 1,101[c] n (%) |
|---|---|---|---|---|---|---|
| | Relevant psychiatric AEs/100PY[b] | 9.6 | All 4 CFTR modulators: 6.9 | | | |
| | | | 9.4 | 5.7 | 5.3 | 5.8 |
| | Participants with relevant psychiatric AE [a] | 64 (3.4) | 263 (12.9) | 144 (7.8) | 149 (8.4) | 57 (5.2) |
| **Number of participants with psychiatric events, N1** | | **N1=64 n/N1, %** | **N1=263 n/N1, %** | **N1=144 n/N1, %** | **N1=149 n/N1, %** | **N1=57 n/N1, %** |
| Seriousness | Serious | 4 (6.3) | 14 (5.3) | 14 (9.7) | 7 (4.7) | 6 (10.5) |
| | Non-serious | 60 (93.8) | 249 (94.7) | 130 (90.3) | 142 (95.3) | 51 (89.5) |
| | Fatal | 0 | 0 | 0 | 0 | 0 |
| Age | Aged <18 years | 13 (20.3) | 64 (24.3) | 27 (18.8) | 35 (23.5) | 15 (26.3) |
| | Aged ≥18 years | 51 (79.7) | 199 (75.7) | 117 (81.3) | 114 (76.5) | 42 (73.7) |
| Sex | Male | 24 (37.5) | 113 (43.0) | 49 (34.0) | 54 (36.2) | 25 (43.9) |
| | Female | 40 (62.5) | 150 (57.0) | 95 (66.0) | 95 (63.8) | 32 (56.1) |
| Past medical history of psychiatric condition | Yes | 25 (39.1) | 125 (47.5) | 66 (45.8) | 56 (37.6) | 22 (38.6) |
| | No | 39 (60.9) | 138 (52.5) | 78 (54.2) | 93 (62.4) | 35 (61.4) |

AE: adverse event; CFTRm: cystic fibrosis transmembrane conductance regulator modulators; ELX: elexacaftor; IVA: ivacaftor; LUM: lumacaftor; n is the number of participants reported per category; N is number of participants treated per product; PT: preferred term; TE: treatment emergent; TEZ: tezacaftor

[a] Relevant Psychiatric PTs: Abnormal behaviour, Anxiety, Completed Suicide, Depression, Depressed mood, Depression suicidal, Emotional disorder, Insomnia, Mental disorder, Mood altered, Sleep disorder, Suicide attempt, Suicidal behaviour, Suicidal ideation, and Suicide threat.

[b] Events/100PY: number of events per 100 patient years (336 days = 48 weeks per year) = number of events / (total duration of cumulative treatment-emergent period / (336*100)).

[c] Participants exposed to multiple treatments are included in the safety sets for each treatment. Participants treated with multiple modulator treatments in multiple treatment arms or open-label extension trials are counted once under each modulator.

- Included all participants with CF aged 1 month and older who received at least 1 dose of study drug (either placebo or in approved dose) from Phase 2 and 3 controlled and uncontrolled trials in IVA, LUM/IVA, TEZ/IVA, and ELX/TEZ/IVA programs. For LUM/IVA Studies 809-102/103/104/105, participants on the 2 doses evaluated in the pivotal Phase 3 trials or placebo in the study safety sets are included.
- A participant is considered in the serious category if there is at least 1 serious event.
- Age and sex are based on the initial exposure of the corresponding treatment.
- Past medical history of psychiatric condition is identified as having any event in the System Organ Class (SOC) of Psychiatric disorders, in the medical history of the earliest study associated with the treatment of the participants.
- AEs are all treatment emergent, defined as AEs started during a cumulative treatment emergent (TE) period, which is defined as the time frame that the participant did not switch to a different treatment and did not have a gap of study participation (e.g., gaps between Phase 2 and Phase 3 studies, or protocol defined wash-out period). The beginning of a cumulative TE period is the date of initial dose of the treatment, and the end of a cumulative TE period is the end of the last TE period (as defined in individual study) within this cumulative TE period.
- VNZ was excluded from these analyses as the pivotal trial included an active comparator without placebo.

**Table E4. Summary of results from 21 studies identified via systematic literature review**

| Study (author, year) | Patients[a] | Follow-up post-treatment initiation | Score Change Post-treatment | | | |
|---|---|---|---|---|---|---|
| | | | Depression | Anxiety | Sleep | Other |
| Allgood, 2023 (2) | 22 adults, US | 14, 28, 42, 56, 70, 84, 98 days | No change in PHQ-8<br>BL mean: 4.9<br>FU mean: 2.9 to 4.3<br>P-value: not significant (value not reported) | No change in GAD-7<br>BL mean: 4.1<br>FU mean: 2.5 to 3.3<br>P-value: not significant (value not reported) | Improvement in PSQI sleep quality<br>BL mean: 7.1,<br>FU mean: 4.6 to 6.2<br>P-value: significant (value not reported) | |
| DiMango, 2021 (3) | 43 adults, US | 3 months | | | Improvement in SNOT-22 sleep domain<br>BL mean: 6.8<br>FU mean: 5.1<br>P-value: <0.05 | Improvement in SNOT-22 psychologic domain<br>BL mean: 11.8<br>FU mean: 9.3<br>P-value: <0.05<br>No change in CFQ-R emotion domain BL mean: 73.3<br>FU mean: 76.0<br>P-value: ≥0.05 |
| Douglas, 2021 (4) | 25 adults, US | 5 months | | | No change in SNOT-22 sleep domain<br>BL mean: 8.6<br>FU mean: 8.1<br>P-value: 0.58 | No change in SNOT-22 psychologic domain<br>BL mean: 8.1<br>FU mean: 6<br>P-value: 0.06 |
| Douglas, 2025 (5) | 108 children (age 6-11 years), Australia | 1 month | | | No change in total SDSC<br>Median change from BL: +1<br>P-value: 0.97 | No change in PSC-17<br>Median change from BL: 0<br>P-value: 0.61 |

| Study (author, year) | Patients[a] | Follow-up post-treatment initiation | Score Change Post-treatment | | | |
|---|---|---|---|---|---|---|
| | | | Depression | Anxiety | Sleep | Other |
| Francesca 2025 (6) | 181 adults and children, Italy | 12, 24, 36 months | | | | Improvement in CFQ-R emotion domain<br>BL mean: 73.3<br>FU mean: 80.0 to 86.7<br>P-value: <0.05 |
| Garcia, 2025 (7) | 108 adults, Spain | 3, 6, 12 months | No change in PHQ-9<br>Median change from BL: -0.5 to -1.5<br>P-value: 0.14 to 0.19 | No change in GAD-7<br>Median change from BL: -1 to -2<br>P-value: 0.09 to 0.43 | | Improvement in CFQ-R emotion domain<br>Median change from BL: 0 to 13.33<br>P-value: <0.001 to 0.02 |
| Graziano, 2024 (8) | 92 adults and children, Italy | 1, 3, 6 months | Improvement in PHQ-9<br>BL mean: 3.7<br>FU mean: 2.7 to 2.9<br>P-value: 0.001 | No change in GAD-7<br>BL mean: 3.7,<br>FU mean: 3.1 to 3.6<br>P-value: 0.37 | | No change in CFQ-R emotion domain<br>BL mean: 80.1<br>FU mean: 83.6 to 84.4<br>P-value: 0.15 |
| Gruber, 2023 (9) | 21 adults, Germany | Mean 5.6 ± 0.8 years | | | | Improvement in CFQ-R emotion domain<br>BL mean: 75.9<br>FU mean: 87.1<br>P-value: 0.046 |
| Hevilla, 2024 (10) | 31 adults, Spain | 12 months | No change in HADS-D<br>BL mean: 2.9<br>FU mean: 2.5<br>P-value: 0.52 | No change in HADS-A<br>BL mean: 4.9<br>FU mean: 4.6<br>P-value: 0.64 | | No change in CFQ-R emotion domain<br>BL mean: 84.5<br>FU mean: 84.3<br>P-value: 0.94 |

| Study (author, year) | Patients[a] | Follow-up post-treatment initiation | Score Change Post-treatment | | | |
|---|---|---|---|---|---|---|
| | | | Depression | Anxiety | Sleep | Other |
| Kos, 2022 (11) | 20 adults and children, Netherlands | 12 months | | | | No change in CFQ-R emotion domain<br>BL mean: 67.9<br>FU mean: 76.1<br>P-value: 0.16 |
| McCoy, 2023 (12) | 18 adults and children, US | 1, 3, 6, 12, 24 months | | | | No change in CFQ-R emotion domain<br>BL median: 73.3<br>FU median: 80.0 to 86.7<br>P-value: 0.17 |
| Nguyen, 2025 (13) | 100 adults, Canada | 6, 12 months | Improvement in PHQ-9<br>Mean change from BL: -1<br>P-value: <0.001 | Improvement in GAD-7<br>Mean change from BL: -1<br>P-value: 0.03 | | |
| Pasley, 2025 (14) | 81 children, US | 1, 3, 6, 9, 12, 18 months | No change in PHQ-8<br>Mean change from BL: -1.3<br>P-value: 0.13<br>Improvement in PROMIS-D<br>Mean change from BL : -2.3 to -3.4<br>P-value: <0.01 to 0.04 | Improvement in GAD-7<br>Mean change from BL: -1.2 to -1.4<br>P-value: 0.01 to 0.04<br>Improvement in PROMIS-A<br>Median BL: 41<br>Median FU: decreased, value not reported<br>P-value: significant, value not reported | | |
| Perez-Ruiz, 2025 (15) | 28 children, | 6 to 8 months | | | | Improvement in CFQ-R emotion domain |

| Study (author, year) | Patients[a] | Follow-up post-treatment initiation | Score Change Post-treatment | | | |
|---|---|---|---|---|---|---|
| | | | Depression | Anxiety | Sleep | Other |
| | Spain | | | | | Mean BL: 78.0<br>Mean FU: 84.0<br>P-value: 0.005 |
| Pham, 2024 (16) | 31 children, Australia | 18 months | No change in PHQ-9<br>BL mean: 6.4<br>FU mean: 6.6<br>Change from BL: 0.24<br>P-value: 0.83 | No change in GAD-7<br>BL mean: 5.3<br>FU mean: 5.9<br>Change from BL: 0.52<br>P-value: 0.66 | No change in SDSC, sleep disturbance<br>BL mean: 43<br>FU mean: 42.9<br>Change from BL: -0.1<br>P-value: 0.96<br>No change in PDSS, sleep disturbance<br>BL mean: 11.5<br>FU mean: 11.7<br>Change from BL: 0.2<br>P-value: 0.85 | |
| Piehler, 2023 (17) | 70 adults, Germany | 8 to 16 weeks | Improvement in PHQ-9<br>Median change from BL: -1<br>P-value: 0.05<br>Improvement in BDI-FS<br>BL median: 1<br>FU median: 0<br>P-value: <0.05 | No change in GAD-7<br>Median BL: 2.0<br>Median FU: 2.0<br>P-value: 0.11 | | No change in CFQ-R emotion domain<br>BL median: 80<br>FU median: 80<br>P-value: 0.37 |
| Pudukodu, 2024 (18) | 86 adults, US | 6 months | No change in PHQ-9<br>BL mean: 5<br>FU mean: 4.8<br>P-value 0.97 | No change in GAD-7<br>BL mean: 4.9<br>FU mean: 5.3<br>P-value: 0.41 | | |

| Study (author, year) | Patients[a] | Follow-up post-treatment initiation | Score Change Post-treatment | | | |
|---|---|---|---|---|---|---|
| | | | Depression | Anxiety | Sleep | Other |
| Sakon, 2023 (19) | 56 adults, US | 3 to 6 months | No change in PHQ-9<br>Median change from BL: -1.11<br>P-value: not reported | | | |
| Vincken, 2025 (20) | 79 adults, Belgium | 3, 6 months | No change in PHQ-9<br>BL median: 4<br>FU median: 3<br>P-value: >0.05 | No change in GAD-7<br>BL median: 3<br>FU median: 2-3<br>P-value: >0.05 | | |
| Welsner, 2022 (21) | 29 adults, Germany | Mean 194 ± 21 days | | | No change or improvement across PSG indices<br>Change from BL: varied<br>P-value: <0.001 to 0.76 | |
| Zhang, 2022 (22) | 100 adults, US | Not reported | No change in PHQ-9<br>BL mean: 5,<br>FU mean: 5.1<br>P-value: 0.9 | No change in GAD-7<br>BL mean: 4.3,<br>FU mean: 4.5<br>P-value: 0.8 | | |

Note: Studies organized alphabetically by last name of first author.

[a] The total number of patients in the study was provided where available; sample size for each outcome based on the number of patients with non-missing data. Information about inclusion of adults, pediatrics, or both are specified where provided in the publication.

BL: baseline; FU: follow-up; BDI-FS: Beck Depression Inventory-Fast Screen; CFQ-R: Cystic Fibrosis Questionnaire-Revised; GAD-7: Generalized Anxiety Disorder 7-item scale; HADS-A: The Hospital Anxiety and Anxiety Scale ; HADS-D: The Hospital Anxiety and Depression Scale ; PDSS: The Pediatric Daytime Sleepiness Scale; PHQ-8: The Patient Health Questionnaire-8; PHQ-9: The Patient Health Questionnaire-9; PROMIS-A: PROMIS

Anxiety; PROMIS-D: PROMIS Depression; PSC-17: Pediatric Symptom Checklist-17; PSG: polysonography; PSQI: Pittsburgh Sleep Quality Index; SDSC: The Sleep Disorder Scale for Children; SNOT-22: The Sino-Nasal Outcome Test.

**Table E5. Results of phenome-wide human genetics association tests in biobanks**

| Study | Variant used | Phenotype | Reference (23-25) | Cases | Controls | beta | P-Value |
|---|---|---|---|---|---|---|---|
| PTV burden | PTV burden | Glycated haemoglobin (HbA1c) | Genebass | 376931 | | -4.41e-03 | 2.38e-07 |
| | PTV burden | Staphylococcus infections | All by all | 1668 | 167839 | 4.83e+00 | 1.35e-06 |
| | PTV burden | Staphylococcus aureus | All by all | 1215 | 169867 | 4.58e+00 | 4.68e-06 |
| | PTV burden | Staphylococcus | All by all | 1480 | 169273 | 4.45e+00 | 8.59e-06 |
| | PTV burden | Mucolytics | All by all | 2252 | 167711 | 4.32e+00 | 1.55e-05 |
| | PTV burden | Methicillin resistant Staphylococcus aureus | All by all | 1504 | 169800 | 4.27e+00 | 1.96e-05 |
| | PTV burden | Drug resistant microorganisms | All by all | 1918 | 168804 | 4.17e+00 | 3.04e-05 |
| | PTV burden | Vitamin K | All by all | 2489 | 167474 | 4.10e+00 | 4.14e-05 |
| | PTV burden | Pneumonia | All by all | 6683 | 155193 | 4.09e+00 | 4.33e-05 |
| | PTV burden | Bronchiectasis | All by all | 736 | 98332 | 3.90e+00 | 9.64e-05 |
| F508del GWAS | F508del | Aspergillosis | FinnGen/UKB | 643 | 869345 | 1.05e+00 | 7.41e-08 |
| | F508del | Chronic pancreatitis | FinnGen/UKB | 5717 | 824949 | 5.88e-01 | 3.43e-07 |

PTV burden tests reported in this table are limited to associations with at least suggestive PTV burden p-value (1e-4). F508del and rs10255092 GWAS results are limited to results associated with at least GWAS suggestive p-value (1e-5).

**Table E6. Results of human genetics association tests with 23 neuropsychiatric traits**

| Study | Variant used | Phenotype | Reference (23, 24, 26-50) | Cases | Controls | beta | P-Value |
|---|---|---|---|---|---|---|---|
| PTV burden | PTV burden | Neuroticism | Genebass | 320534 | | 1.98e-04 | 8.43e-01 |
| | PTV burden | Schizophrenia | SCHEMA | 24248 | 97322 | -1.59e-01 | 6.01e-01 |
| | PTV burden | Anxiety | Genebass | 18338 | 376503 | 2.43e-03 | 5.83e-01 |
| | PTV burden | Bipolar | BipEx | 13933 | 14422 | -1.06e+00 | 1.47e-01 |
| | PTV burden | Substance addiction and disorders | All by all | 10827 | 153215 | -4.28e-01 | 6.69e-01 |
| | PTV burden | Depression | Tian, 2024 | 10800 | 256930 | -3.59e-01 | 4.55e-01 |
| | PTV burden | Long sleep | Genebass | 10225 | 39517 | -4.75e-03 | 4.56e-01 |
| | PTV burden | Insomnia | All by all | 8175 | 149948 | -3.20e-01 | 7.49e-01 |
| | PTV burden | Autism | ASC | 5556 | 8809 | | 8.40e-01 |
| | PTV burden | Attention-deficit hyperactivity disorders | All by all | 3098 | 167976 | -8.02e-01 | 4.23e-01 |
| | PTV burden | Suicide attempt | Genebass | 2830 | 2489 | -1.32e-02 | 3.78e-01 |
| | PTV burden | Posttraumatic stress disorder | All by all | 2806 | 164364 | 1.07e+00 | 2.85e-01 |

| | | | | | | | |
|---|---|---|---|---|---|---|---|
| | PTV burden | Parkinson's | Genebass | 1901 | 392940 | 3.51e-03 | 7.90e-01 |
| | PTV burden | Alzheimer's | Genebass | 900 | 393941 | -1.18e-02 | 5.38e-01 |
| | PTV burden | Eating disorders | Genebass | 681 | 212932 | -2.82e-02 | 2.05e-01 |
| | PTV burden | Obsessive-compulsive disorders | Genebass | 660 | 394181 | -1.98e-03 | 9.26e-01 |
| F508del GWAS | rs113827944 | Neuroticism | Nagel, 2018* | 390278 | | 3.32e-04 | 9.68e-01 |
| | rs113827944 | Worry | Nagel, 2018 | 348219 | | 1.09e-02 | 2.42e-01 |
| | F508del | Antidepressant use | Levey, 2025 | 191800 | 586152 | -4.39e-02 | 3.72e-01 |
| | rs113827944 | PTSD | Nievergelt, 2024 | 137136 | 1086146 | 3.77e-03 | 5.90e-01 |
| | F508del | Alcohol use disorder | Zhou, 2023 | 113325 | 639923 | 2.52e-03 | 9.31e-01 |
| | F508del | Insomnia | Watanabe, 2022 | 109548 | 277440 | -1.21e+00 | 2.27e-01 |
| | rs113827944 | Alzheimer's | Wightman, 2021 | 86531 | 676386 | 2.13e-03 | 7.74e-01 |
| | rs113827944 | Schizophrenia | Trubetskoy, 2022 | 53386 | 77258 | -2.46e-02 | 4.96e-01 |
| | F508del | Short sleep | Austin-Zimmerman, 2023 | 47054 | 382950 | -5.80e-03 | 8.54e-01 |
| | F508del | Cannabis use disorder | Levey, 2023 | 42281 | 843744 | 4.55e-02 | 7.72e-01 |
| | rs113827944 | ADHD | Demontis, 2023 | 38691 | 186843 | -3.98e-02 | 3.08e-01 |

| | | | | | | | |
|---|---|---|---|---|---|---|---|
| | rs113827944 | OCS | Strom, 2024 | 33942 | | -9.90e-03 | 7.74e-01 |
| | rs113827944 | Suicide attempt | Dochetry, 2023 | 31094 | 758690 | 2.08e-02 | 5.80e-01 |
| | F508del | Autism | Grove, 2019 | 18381 | 27969 | 1.00e-01 | 1.10e-01 |
| | F508del | Anorexia nervosa | Watson, 2019 | 16992 | 55525 | -2.06e-02 | 7.37e-01 |
| | F508del | Anxiety | Neale, 2018 | 16730 | 101021 | 1.26e-02 | 2.59e-02 |
| | F508del | Long sleep | Austin-Zimmerman, 2023 | 15962 | 382950 | 4.29e-02 | 3.95e-01 |
| | rs113827944 | Opioid use disorder | Deak, 2022 | 15251 | 538935 | 7.95e-02 | 2.42e-03 |
| | rs113827944 | Parkinson's | Nalls, 2019 | 15056 | 12637 | -4.72e-02 | 6.71e-01 |
| | F508del | Bipolar disorder | BipEx | 14210 | 14422 | 3.37e-02 | 6.39e-01 |
| | rs113827944 | Tourette | Yu, 2018 | 4819 | 9488 | -1.84e-01 | 4.95e-02 |
| | rs113827944 | OCD | Arnold, 2018 | 2688 | 7037 | -3.93e-02 | 7.39e-01 |
| | rs113827944 | Panic disorder | Forstner, 2021 | 2248 | 7992 | 1.93e-02 | 9.01e-01 |
| | rs10255092 | Neuroticism | Nagel, 2018* | 390278 | | 4.79e-03 | 6.28e-02 |
| rs10255092 GWAS | rs10255092 | Worry | Nagel, 2018 | 348219 | | -7.10e-03 | 8.58e-03 |
| | rs10255092 | Antidepressant use | Levey, 2025 | 191800 | 586152 | 7.30e-03 | 1.30e-01 |

| | | | | | | | |
|---|---|---|---|---|---|---|---|
| | rs10255092 | PTSD | Nievergelt, 2024 | 137136 | 1086146 | 9.80e-04 | 6.15e-01 |
| | rs10255092 | Alcohol use disorder | Zhou, 2023 | 113325 | 639923 | 2.32e-03 | 3.84e-01 |
| | rs10255092 | Insomnia | Watanabe, 2022 | 109548 | 277440 | -1.21e+00 | 2.27e-01 |
| | rs10255092 | Alzheimer's | Wightman, 2021 | 86531 | 676386 | 2.65e-04 | 8.79e-01 |
| | rs10255092 | Bipolar disorder | O'Connell, 2025 | 59287 | 781022 | -8.00e-04 | 9.20e-01 |
| | rs10255092 | Schizophrenia | Trubetskoy, 2022 | 53386 | 77258 | -2.01e-02 | 3.57e-02 |
| | rs10255092 | Short sleep | Austin-Zimmerman, 2023 | 47054 | 382950 | 7.20e-03 | 3.71e-01 |
| | rs10255092 | Cannabis use disorder | Levey, 2023 | 42281 | 843744 | 2.50e-03 | 7.97e-01 |
| | rs10255092 | ADHD | Demontis, 2023 | 38691 | 186843 | -4.70e-03 | 6.48e-01 |
| | rs10255092 | OCS | Strom, 2024 | 33942 | | 4.30e-03 | 6.18e-01 |
| | rs10255092 | Suicide attempt | Dochetry, 2023 | 31094 | 758690 | -8.40e-03 | 3.90e-01 |
| | rs10255092 | Autism | Grove, 2019 | 18381 | 27969 | 7.30e-03 | 6.33e-01 |
| | rs10255092 | Anorexia nervosa | Watson, 2019 | 16992 | 55525 | 1.74e-02 | 2.73e-01 |
| | rs10255092 | Anxiety | Neale, 2018 | 16730 | 101021 | 1.79e-03 | 2.69e-01 |
| | rs10255092 | Long sleep | Austin-Zimmerman, 2023 | 15962 | 382950 | 1.41e-02 | 2.86e-01 |

| | | | | | | | |
|---|---|---|---|---|---|---|---|
| | rs10255092 | Opioid use disorder | Deak, 2022 | 15251 | 538935 | -1.23e-03 | 8.47e-01 |
| | rs10255092 | Parkinson's | Nalls, 2019 | 15056 | 12637 | -2.96e-02 | 2.02e-01 |
| | rs10255092 | Tourette | Yu, 2018 | 4819 | 9488 | 3.24e-02 | 2.69e-01 |
| | rs10255092 | OCD | Arnold, 2018 | 2688 | 7037 | 3.58e-02 | 3.44e-01 |
| | rs10255092 | Panic disorder | Forstner, 2021 | 2248 | 7992 | 6.30e-03 | 8.72e-01 |

* Results obtained from the NDKP browser (https://ndkp.hugeamp.org/, accessed Sep 19th, 2025)

7. Garcia MS, Pelaez A, Punter RMG, Lopez MC, Carbajal CM, Ancochea J, Bachiller JME, Hernandez ASM, Rodrigo-Garcia M, Clemente MG, Moreno RMG. Unveiling the

psychosocial impact of Elexacaftor/Tezacaftor/Ivacaftor therapy in Cystic Fibrosis patients. *BMC Pulm Med* 2025; 25: 81.

8. Graziano S, Boldrini F, Pellicano GR, Milo F, Majo F, Cristiani L, Montemitro E, Alghisi F, Bella S, Cutrera R, Fiocchi AG, Quittner A, Tabarini P. Longitudinal Effects of Elexacaftor/Tezacaftor/Ivacaftor: Multidimensional Assessment of Neuropsychological Side Effects and Physical and Mental Health Outcomes in Adolescents and Adults. *Chest* 2024; 165: 800-809.
9. Gruber W, Welsner M, Blosch C, Dillenhoefer S, Olivier M, Brinkmann F, Koerner-Rettberg C, Sutharsan S, Mellies U, Taube C, Stehling F. Long-Term Follow-Up of Health-Related Quality of Life and Short-Term Intervention with CFTR Modulator Therapy in Adults with Cystic Fibrosis: Evaluation of Changes over Several Years with or without 33 Weeks of CFTR Modulator Therapy. *Healthcare (Basel)* 2023; 11.
10. Hevilla F, Porras N, Giron MV, Garcia-Olivares M, Padial M, Sanchez-Torralvo FJ, Olveira C, Olveira G. Impact of Elexacaftor-Tezacaftor-Ivacaftor Therapy on Body Composition, Dietary Intake, Biomarkers, and Quality of Life in People with Cystic Fibrosis: A Prospective Observational Study. *Nutrients* 2024; 16.
11. Kos R, Neerincx AH, Fenn DW, Brinkman P, Lub R, Vonk SEM, Roukema J, Reijers MH, Terheggen-Lagro SWJ, Altenburg J, Majoor CJ, Bos LD, Haarman EG, Maitland-van der Zee AH, Amsterdam Mucociliary Clearance Disease Research G. Real-life efficacy and safety of elexacaftor/tezacaftor/ivacaftor on severe cystic fibrosis lung disease patients. *Pharmacol Res Perspect* 2022; 10: e01015.

30. Deak JD, Zhou H, Galimberti M, Levey DF, Wendt FR, Sanchez-Roige S, et al. Genome-wide association study in individuals of European and African ancestry and multi-trait

analysis of opioid use disorder identifies 19 independent genome-wide significant risk loci. Mol Psychiatry. 2022;27(10):3970–9.
